# Recurrent Single-Nucleotide Insertions in the Mitochondrial Second Light-Strand Promoter Cause Tubulointerstitial Kidney Disease

**DOI:** 10.64898/2026.07.31.26359118

**Authors:** Klára Svojšová, Kendrah O. Kidd, Dita Mušálková, Tereza Kmochová, Hana Hartmannová, Kateřina Hodaňová, Viktor Stránecký, Veronika Barešová, Helena Trešlová, Martin Radina, Lea Pavlovičová, Jakub Sikora, Abbigail Taylor, Lauren Martin, Antonio Sanchez, Nelson Weller, Thomas Pinder, Megan E. Astley, Xiangling Wang, Abhijit Dixit, Stephen M. Korbet, Colm Rowan, Peter J Conlon, Karina Soto, Afonso Santos, Marek Mysliveček, Jiří Zeman, Hana Štufková, Hana Hansíková, Kateřina Tauchmannová, Petr Pecina, Vilma Kaplanová, Marek Vrbacký, Tomáš Mráček, Örjan Persson, Claes M. Gustafsson, Maria Falkenberg, Martina Živná, Anthony J. Bleyer, Stanislav Kmoch

## Abstract

**Introduction:** Mitochondrial DNA (mtDNA) is not routinely analyzed in inherited kidney disease. We evaluated mtDNA variation in families who remained genetically unresolved despite extensive testing.

**Methods:** We reviewed pedigrees from the Wake Forest–Charles University Rare Inherited Kidney Disease Registry to identify genetically unresolved families with suspected maternal inheritance, performed mtDNA genotyping, clinically characterized variant carriers, and functionally evaluated disease-associated mitochondrial variants.

**Results:** Among 33 families with evidence of maternal inheritance, 18 (55%) carried one of seven disease-associated mtDNA variant types, including homoplasmic recurrent single-nucleotide insertions in the second light-strand promoter (LSP2; 9 families), novel *MT-TW* and *MT-TL2* variants (2 and 1 families, respectively), and previously reported *MT-TF* and heteroplasmic *MT-ND5* variant (5 and 1 families, respectively). In 16 families, variants occurred on distinct haplotypes, consistent with independent mutational events and rapid enrichment to homoplasmy across generations. Maternal transmission was strongly supported, with below-normal kidney function observed in 54/60 (90%) offspring of affected mothers versus 1/17 (6%) offspring of affected fathers (p = 1.23 × 10^⁻11^). Pathogenicity was further supported by predicted deleterious structural effects and functional evidence of impaired mitochondrial transcription and translation, respiratory chain deficiency, and CoQ10 depletion. Affected individuals predominantly presented with chronic tubulointerstitial kidney disease, occasionally accompanied by gout and only sporadically with extrarenal manifestations. The rate of kidney disease progression appeared to vary both between and within families. Overall, 109/119 genetically affected individuals or obligate at-risk carriers were clinically affected; most unaffected carriers were younger than 45 years of age. Clinical status was unavailable for an additional 66 obligate at-risk carriers.

**Conclusions:** These findings establish the physiological relevance of the LSP2 promoter, support routine assessment of the mitochondrial genome in inherited kidney disease, and highlight mtDNA variants as an important cause of familial and sporadic tubulointerstitial kidney disease of previously unexplained etiology.

**Lay Summary:** Many inherited kidney diseases remain unexplained because routine genetic testing focuses on genes in the cell nucleus and does not examine mitochondrial DNA—the small genome in the cell’s energy-providing mitochondria, inherited only from the mother. We studied 33 families with chronic kidney disease whose family histories suggested maternal inheritance and identified disease-causing mitochondrial DNA variants in 18 (55%). Nine families carried variants in LSP2, a recently discovered mitochondrial regulatory element, highlighting its importance in normal mitochondrial function and disease. Others carried pathogenic variants in mitochondrial tRNA genes required for mitochondrial protein synthesis. Laboratory studies showed that these variants impair mitochondrial energy conversion. In all families, the predominant manifestation was slowly progressive kidney disease, sometimes leading to dialysis or kidney transplantation. These findings identify pathogenic mitochondrial DNA variants as an underrecognized cause of inherited kidney disease and support the inclusion of mitochondrial DNA analysis in routine genetic testing.

## Introduction

Hereditary tubulointerstitial kidney disease (TKD) is a genetically heterogeneous group of disorders most commonly characterized by a bland urinary sediment, absence of proteinuria, and progressive chronic kidney disease (CKD), typically leading to kidney failure between the ages of 20 and 80.^1^ The autosomal dominant form (ADTKD) is considered the third most common monogenic kidney disease in adults after autosomal dominant polycystic kidney disease and Alport syndrome.^2, 3^ While causative pathogenic variants underlying ADTKD have been identified in *MUC1*, *UMOD*, *HNF1B*, *REN*, *SEC61A1*, *APOA1*, and *APOA4*, a subset of cases remain genetically unresolved.^4^

Several reports have identified pathogenic variants in mitochondrial DNA as a cause of TKD,^5–10^ leading to the proposal of the term “mitochondrially inherited tubulointerstitial kidney disease” (MITKD).^8^ Mitochondrial disorders should therefore be considered in the differential diagnosis of cases with hereditary TKD of unclear genetic etiology, particularly when maternal inheritance is possible.

Mitochondrial DNA is not routinely assessed by standard diagnostic approaches such as multi-gene panels, whole-exome sequencing (WES), or whole-genome sequencing (WGS) in cases of inherited kidney disease. To address this gap, we evaluated mitochondrial DNA variation in 33 families from the Wake Forest-Charles University Rare Inherited Kidney Disease Registry (WF-CUNI-RIKD registry) who had previously tested negative in genetic analyses focused on autosomal dominant inheritance, but who upon clinical re-evaluation showed evidence consistent with maternal inheritance. We report the genetic, clinical, structural, and biochemical correlates of seven types of mitochondrial disease-associated variants identified in 18 families (55%), four of which are novel. These include two recurrent single-nucleotide insertions in the recently identified mitochondrial second light-strand promoter (LSP2)^11^ observed in nine families, a unique mitochondrial *MT-TW* variant in two families, and a rare variant in mitochondrial *MT-TL2* in one family, along with three inherited kidney disease-associated variant types in *MT-TF* and *MT-ND5* that have been previously reported.^6, 9, 10^

## Methods

### Ethical approval

This study was approved by the Institutional Review Boards of the First Faculty of Medicine, Charles University in Prague and the Wake Forest University Health Science Institutional Review Board. It adhered to the Declaration of Helsinki. Patients provided informed consent.

### Clinical evaluation and study population

The WF-CUNI-RIKD registry is comprised of over 1400 families referred to A.J.B. by physicians and/or family members since 1996.^12, 13^ A systematic review of all families with inherited tubulointerstitial kidney disease was undertaken to evaluate for mitochondrial inheritance.

### Genetic investigations

Genomic DNA was extracted from whole blood, hair follicles, buccal swabs, or patient-derived fibroblasts using standard procedures. Genomic sequencing, data analysis, variant calling and variant prioritization are described in the **Supplementary Methods**. Heteroplasmy levels were determined from sequencing data as the proportion of mitochondrial reads carrying the variant relative to the total number of reads at the corresponding position.

For pedigree construction and clinical description, the following terminology was used: genetically affected individuals were genetically tested and confirmed to carry the variant; obligate at-risk carriers were untested relatives presumed to carry the variant based on maternal inheritance; and clinically affected individuals had an estimate glomerular filtration rate (eGFR) more than 2 standard deviations below the age- and sex-adjusted means.^14^ Individuals with unknown clinical status had no available serum creatinine measurements. P-values for maternal inheritance were calculated using a two proportion Z-test.

### Molecular and structural correlates of disease-associated LSP2 and tRNA variants

Disease-associated variants in LSP2 were mapped to the revised Cambridge Reference Sequence and analyzed in the context of predicted binding sites of core mitochondrial DNA transcription machinery components, including mitochondrial transcription factor A (TFAM), mitochondrial transcription factor B2 (TFB2M), and mitochondrial RNA polymerase (POLRMT).^11^ Conservation of LSP spacer sequence lengths^11^ was assessed across humans and three primate species retrieved from GenBank using the ClustalW program.^15^ Disease-associated variants in the mitochondrial tRNA genes (*MT-TF*, *MT-TW* and *MT-TL2*) were mapped onto their predicted secondary structures, which were retrieved from MitoVisualize.^16^ The positions of post-transcriptional modifications of the corresponding tRNAs were obtained from Suzuki et al.^17^

### *In vitro* transcription from synthetic heavy- and light-strand mitochondrial promoters

Synthetic DNA templates containing either HSP, LSP2, or both promoters in their native orientation were cloned into the pEX-A128 vector. Corresponding patient-derived constructs carrying single-nucleotide insertions (+T or +G) and additional dual-promoter constructs with −2 to +5 nt insertions in either promoter were generated to assess spacing effects between promoter elements and the TFAM-binding site. Linear templates for run-off transcription assays were produced by PCR amplification from plasmids and analyzed as previously described.^18, 19^ For dual-promoter templates, LSP2 transcription was quantified and normalized to HSP transcription within the same reaction. Statistical analysis was performed using one-way ANOVA with Tukey’s multiple comparisons test in GraphPad Prism.

### Patient-derived materials

Skin fibroblasts were obtained and maintained as previously described.^20^

### Metabolic pulse–chase labeling of mtDNA-encoded proteins

mtDNA-encoded proteins were labeled using ^35^S-Protein Labelling Mix (Met+Cys; Revvity NEG072), separated by SDS-PAGE, and analyzed sequentially by autoradiography and Western blotting as previously described.^21^

### Content of mitochondrial respiratory chain complexes

Separation of native OXPHOS complexes by blue-native electrophoresis (BNE) was performed on polyacrylamide gradient as previously described,^22^ with modifications provided in the **Supplementary Methods**.

### Mitochondrial respiration measurement

The oxygen consumption rate of fibroblast cells was measured at 37 °C using the Oroboros Oxygraph-2k, essentially as previously described,^23^ with modifications provided in the **Supplementary Methods.**

### Total coenzyme Q10 content

Frozen pellets (fibroblasts) were homogenized^24^ and CoQ10 was determined as described.^25^ Citrate synthase (CS) activity was determined according to Srere.^26^

### Label-free quantification mass spectrometry analysis

Label-free quantification mass spectrometry analysis (LFQ-MS) of cell pellets was performed as described.^27^ Protein groups quantities were evaluated in Perseus v. 2.1.5.0.^28^

All methods are described in detail in the **Supplementary Methods**.

## Results

### Genetic investigation

There were 33 genetically unresolved families in the WF-CUNI-RIKD registry with evidence of maternal inheritance. Of the families included in the study, 18 (55%) were found to carry mitochondrial variants that were either unique (i.e., not reported in databases including gnomAD, MITOMAP, HelixMTdb or CzechGenomes) or rare (with a homoplasmic allele count in gnomAD v.3.1.2 of fewer than five of ∼56430 cases). These variants included two types of recurrent single-nucleotide insertions in the recently identified second light-strand promoter (LSP2)^11^ observed in nine families, a unique mitochondrial *MT-TW* variant in two families, a rare variant in mitochondrial *MT-TL2* in one family, and three previously reported MITKD-associated variant types in *MT-TF* and *MT-ND5*^6, 9, 10^ in six additional families (see **Table 1**). In 16 families, disease-associated variants were present on distinct mitochondrial haplotypes, suggesting independent mutational events. In two families with the m.5542C>T variant in *MT-TW*, the variant was present on the identical haplogroup U2e2a1, indicating a distant shared ancestry. Genotyping and low coverage whole-genome sequencing revealed 60 genetically affected individuals, with all disease-associated variant types present in blood at near-complete or complete homoplasmy, except for the *MT-ND5* variant, which was detected at a heteroplasmy level of 5–10%.

**Table 1.**
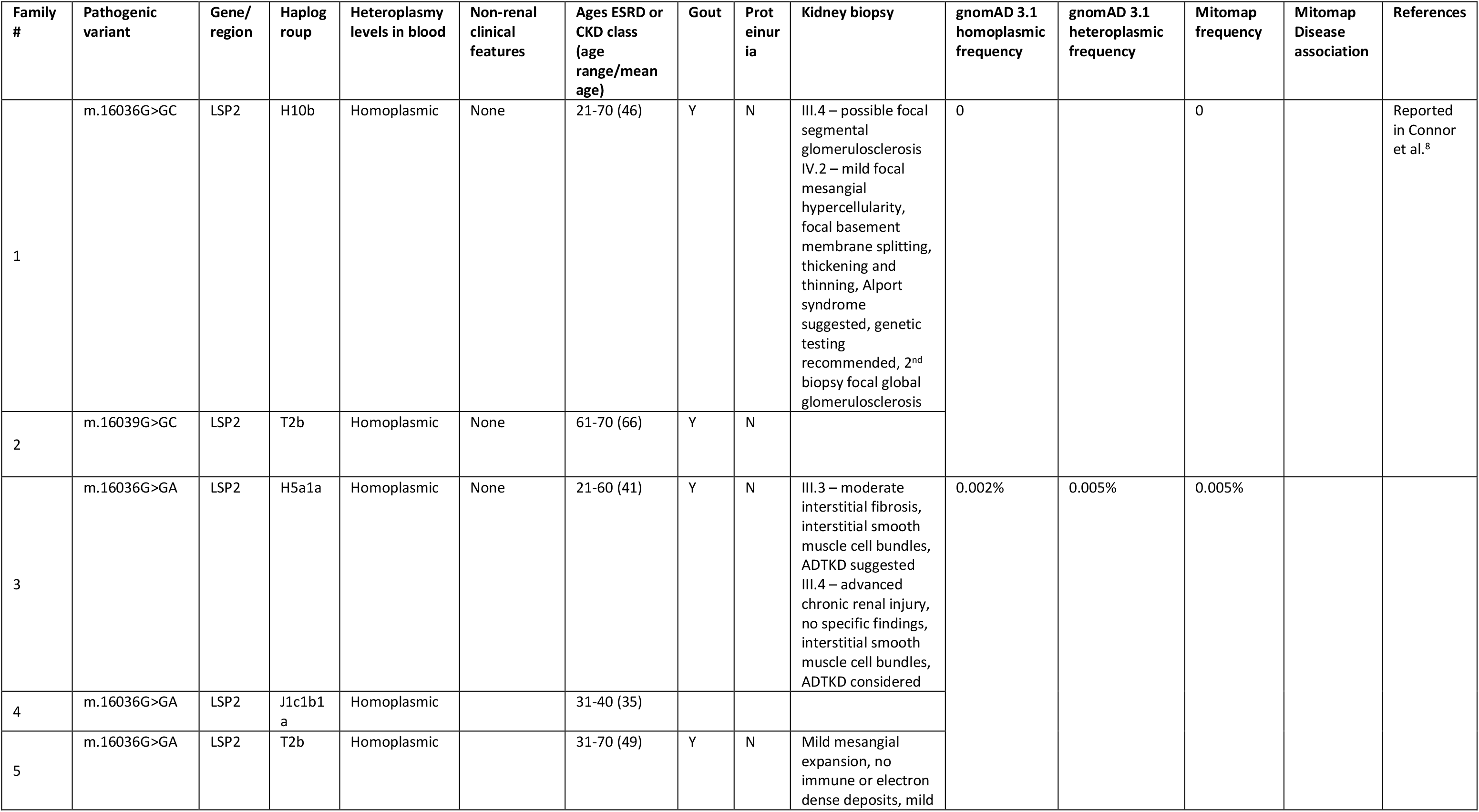
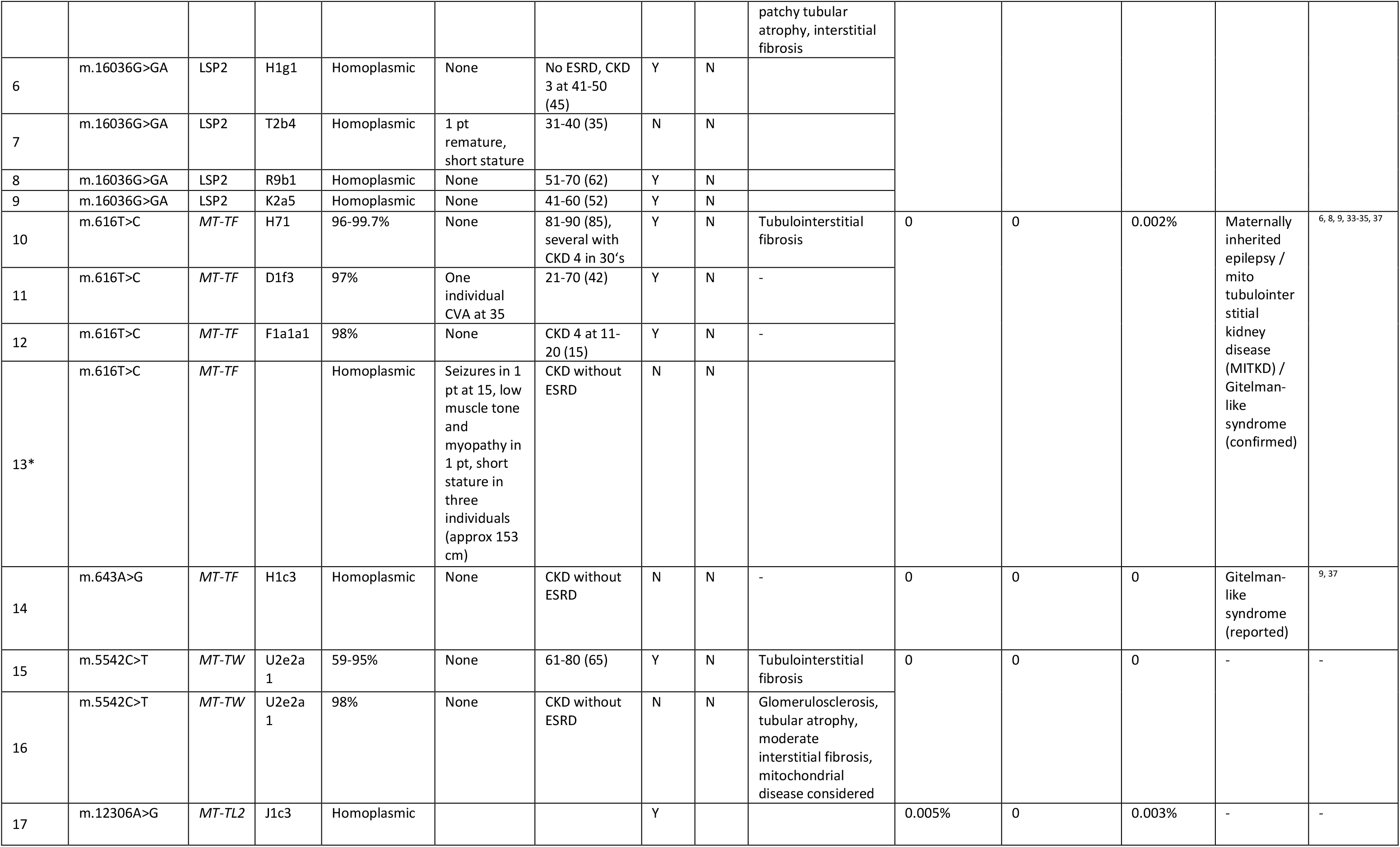
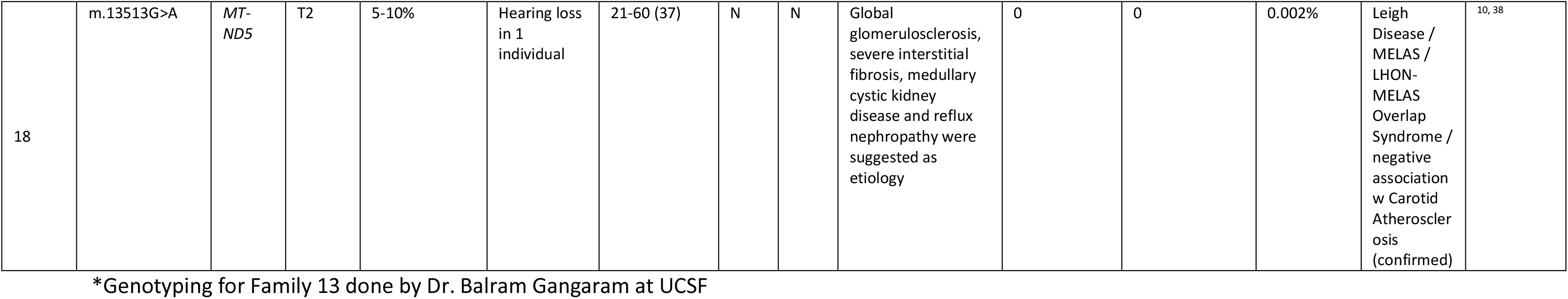
Genetic characteristics of mitochondrial disease-associated variants previously reported and reported in this investigation.

| Family # | Pathogenic variant | Gene/region | Haplogroup | Heteroplasmy levels in blood | Non-renal clinical features | Ages ESRD or CKD class (age range/mean age) | Gout | Proteinuria | Kidney biopsy | gnomAD 3.1 homoplasmic frequency | gnomAD 3.1 heteroplasmic frequency | Mitomap frequency | Mitomap Disease association | References |
| --- | --- | --- | --- | --- | --- | --- | --- | --- | --- | --- | --- | --- | --- | --- |
| 1 | m.16036G>GC | LSP2 | H10b | Homoplasmic | None | 21-70 (46) | Y | N | III.4 – possible focal segmental glomerulosclerosis<br>IV.2 – mild focal mesangial hypercellularity, focal basement membrane splitting, thickening and thinning, Alport syndrome suggested, genetic testing recommended, 2<sup>nd</sup> biopsy focal global glomerulosclerosis | 0 |  | 0 |  | Reported in Connor et al.<sup>8</sup> |
| 2 | m.16039G>GC | LSP2 | T2b | Homoplasmic | None | 61-70 (66) | Y | N |  |  |  |  |  |  |
| 3 | m.16036G>GA | LSP2 | H5a1a | Homoplasmic | None | 21-60 (41) | Y | N | III.3 – moderate interstitial fibrosis, interstitial smooth muscle cell bundles, ADTKD suggested<br>III.4 – advanced chronic renal injury, no specific findings, interstitial smooth muscle cell bundles, ADTKD considered | 0.002% | 0.005% | 0.005% |  |  |
| 4 | m.16036G>GA | LSP2 | J1c1b1a | Homoplasmic |  | 31-40 (35) |  |  |  |  |  |  |  |  |
| 5 | m.16036G>GA | LSP2 | T2b | Homoplasmic |  | 31-70 (49) | Y | N | Mild mesangial expansion, no immune or electron dense deposits, mild patchy tubular atrophy, interstitial fibrosis |  |  |  |  |  |
| 6 | m.16036G>GA | LSP2 | H1g1 | Homoplasmic | None | No ESRD, CKD 3 at 41-50 (45) | Y | N |  |  |  |  |  |  |
| 7 | m.16036G>GA | LSP2 | T2b4 | Homoplasmic | 1 pt remature, short stature | 31-40 (35) | N | N |  |  |  |  |  |  |
| 8 | m.16036G>GA | LSP2 | R9b1 | Homoplasmic | None | 51-70 (62) | Y | N |  |  |  |  |  |  |
| 9 | m.16036G>GA | LSP2 | K2a5 | Homoplasmic | None | 41-60 (52) | Y | N |  |  |  |  |  |  |
| 10 | m.616T>C | MT-TF | H71 | 96-99.7% | None | 81-90 (85), several with CKD 4 in 30's | Y | N | Tubulointerstitial fibrosis | 0 | 0 | 0.002% | Maternally inherited epilepsy / mito tubulointerstitial kidney disease (MITKD) / Gitelman-like syndrome (confirmed) | 6, 8, 9, 33-35, 37 |
| 11 | m.616T>C | MT-TF | D1f3 | 97% | One individual CVA at 35 | 21-70 (42) | Y | N | - |  |  |  |  |  |
| 12 | m.616T>C | MT-TF | F1a1a1 | 98% | None | CKD 4 at 11-20 (15) | Y | N | - |  |  |  |  |  |
| 13* | m.616T>C | MT-TF |  | Homoplasmic | Seizures in 1 pt at 15, low muscle tone and myopathy in 1 pt, short stature in three individuals (approx 153 cm) | CKD without ESRD | N | N |  |  |  |  |  |  |
| 14 | m.643A>G | MT-TF | H1c3 | Homoplasmic | None | CKD without ESRD | N | N | - | 0 | 0 | 0 | Gitelman-like syndrome (reported) | 9, 37 |
| 15 | m.5542C>T | MT-TW | U2e2a1 | 59-95% | None | 61-80 (65) | Y | N | Tubulointerstitial fibrosis | 0 | 0 | 0 | - | - |
| 16 | m.5542C>T | MT-TW | U2e2a1 | 98% | None | CKD without ESRD | N | N | Glomerulosclerosis, tubular atrophy, moderate interstitial fibrosis, mitochondrial disease considered |  |  |  |  |  |
| 17 | m.12306A>G | MT-TL2 | J1c3 | Homoplasmic |  |  | Y |  |  | 0.005% | 0 | 0.003% | - | - |
| 18 | m.13513G>A | MT-ND5 | T2 | 5-10% | Hearing loss in 1 individual | 21-60 (37) | N | N | Global glomerulosclerosis, severe interstitial fibrosis, medullary cystic kidney disease and reflux nephropathy were suggested as etiology | 0 | 0 | 0.002% | Leigh Disease / MELAS / LHON-MELAS Overlap Syndrome / negative association w Carotid Atherosclerosis (confirmed) | 10, 38 |

\*Genotyping for Family 13 done by Dr. Balram Gangaram at UCSF

### Clinical characterization

#### Families with the LSP2 variant

There were nine families identified with LSP2 variants: seven with m.16036G>GA and two with m.16039G>GC. Only one of 12 children of genetically affected fathers had below-normal kidney function, whereas 21 of 22 children of genetically affected mothers were clinically affected (p=2.4 x 10^-7^), supporting mitochondrial inheritance (see **Figure 1**). Moreover, the LSP2 variants were the only variants found in all families, with no other candidate regions or suspicious variants found elsewhere in their nuclear or mitochondrial genomes. The only clinical manifestations of mitochondrial dysfunction were chronic kidney disease and gout (see **Table 1; Supplementary Table S1**). The urinary sediment was normal, and proteinuria was absent.

**Figure 1.**
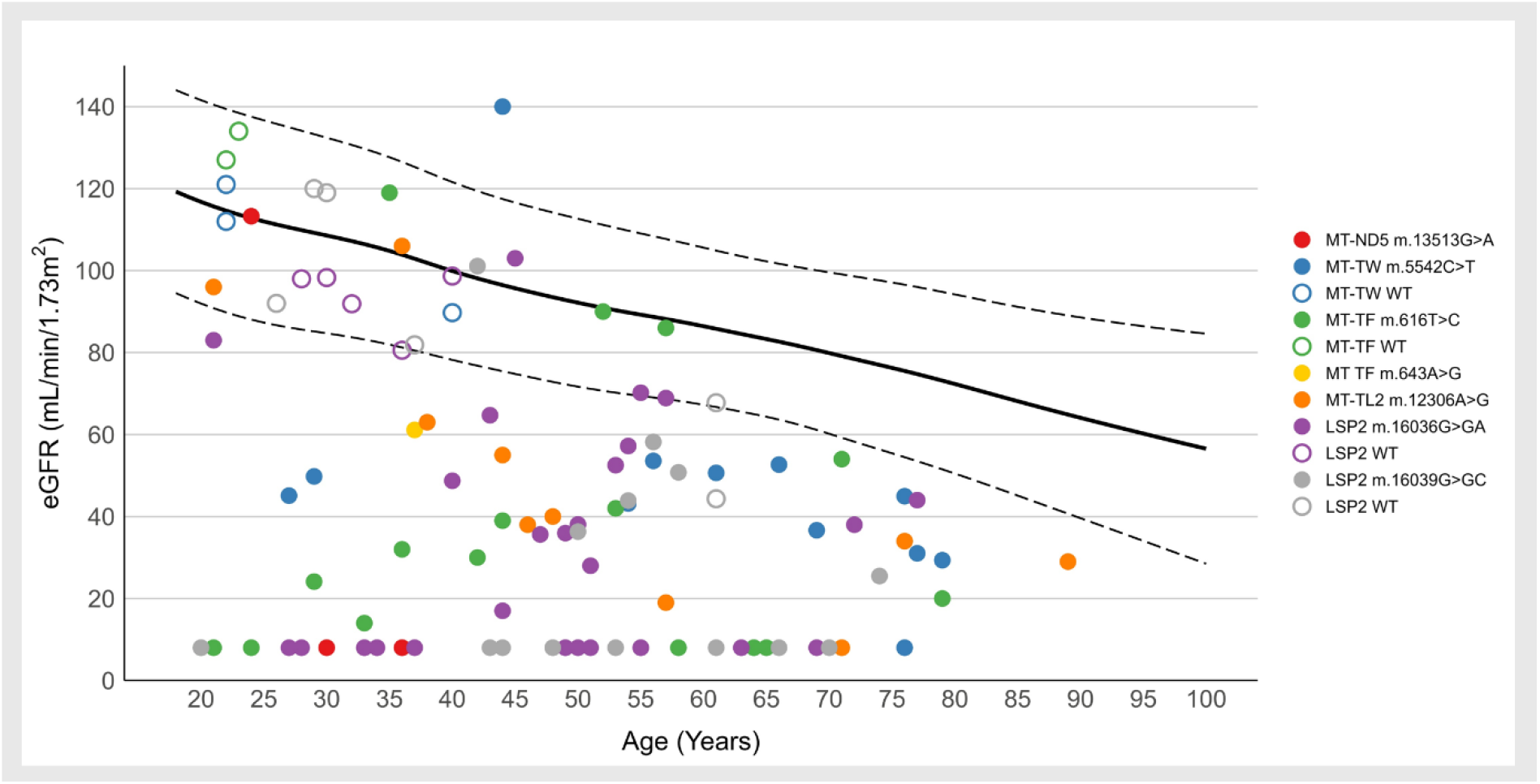
Kidney function in investigated families. The most recent estimated glomerular filtration rate (eGFR, ml/min per 1.73 m^2^) versus age in individuals carrying identified mitochondrial variants and genetically unaffected individuals from investigated families. Individuals on dialysis/transplanted were assigned an eGFR of 8 ml/min per 1.73 m^2^. Full circles denote maternal relatives and thus obligate carriers of the mitochondrial variant. Empty circles denote non-maternal relatives and thus obligate non-carriers from the indicated families. The black line and the two dashed lines represent the mean eGFR ± 2 standard deviation for individuals of western European descent.

#### Families with other mitochondrial variants

Nine families harbored five additional types of mitochondrial variants. Consistent with findings for LSP2 variants, all five children with clinical data of genetically affected fathers had normal kidney function, whereas 33 of 38 children of genetically affected mothers were clinically affected (p =1.5 × 10⁻^5^) (see **Figure 1**), supporting mitochondrial inheritance. No other candidate regions or potentially disease-associated variants were identified elsewhere in the nuclear genome. Patients all suffered from chronic tubulointerstitial kidney disease, with the occasional presence of gout and other clinical manifestations of mitochondrial disease (see **Table 1; Supplementary Table S1**), including short stature, seizures, myopathy and hearing loss. However, these non-renal presentations were not uniform within the families, but rather case-specific.

In summary, 54/60 genetically affected individuals and 55/59 obligate at-risk carriers were clinically affected with reduced age-adjusted eGFR (see **Figure 1**) based on the European Kidney Function Consortium reference values,^14^ while 8 of the remaining 10 individuals had normal kidney function at ages of 20-45, and clinical status was unknown for an additional 66 obligate at-risk carriers at the time of evaluation. The rate of progression of CKD appeared to be variable between families (see **Figure 1**), though there was limited statistical power for evaluation.

### Histopathological findings

A total of eight kidney biopsy reports from seven affected patients were available for review. In six patients, scanned images of routine histological stains (hematoxylin and eosin, Periodic acid-Schiff, trichrome, and methenamine silver) were also available and underwent a second-look assessment. Overall, the histopathological findings were non-specific and non-diagnostic, consisting mainly of varying degrees of interstitial fibrosis, tubular atrophy, and glomerulosclerosis without a distinctive pattern suggestive of a specific disease entity. Immunofluorescence studies were unremarkable. Ultrastructural examination revealed a broad spectrum of changes, most notably variable thickening and thinning of the glomerular basement membrane, but no electron-dense deposits or immune-type structures. Mitochondrial abnormalities were not reported in the original biopsy evaluations; however, re-examination identified swollen mitochondria in proximal tubular cells in two affected individuals. Based on the clinical presentation and family history, ADTKD was considered in three patients, while a mitochondrial disorder was suspected in one family because of maternal inheritance. A summary of the final histopathological diagnoses is provided in **Table 1**; detailed findings are presented in **Supplementary Figure S1**.

### Structural correlates of identified mitochondrial DNA variants

#### LSP2 single-nucleotide insertions and mitochondrial transcriptional architecture

**Figure 2** depicts organization of the mitochondrial genome and critical components of its genome replication, transcription, and their regulation. LSP2 is one of two promoters for light (L)-strand transcription and drives transcription *in vitro*, while influencing steady-state levels of L-strand–derived transcripts *in vivo*; however, its physiological function remains unclear.^29^ Acting independently of replication, it is hypothesized to support high metabolic demands in post-mitotic cells such as kidney, cardiac and skeletal muscle cells, which are particularly susceptible to mitochondrial disease.^30^ The sequence conservancy and organization of LSP2 are depicted in **Figure 3**. The promoter contains a binding site for mitochondrial transcription factor A (TFAM) upstream of the transcription start site, where the polymerase initiation complex consisting of mitochondrial RNA polymerase (POLRMT) and mitochondrial transcription factor B2 (TFB2M) is recruited. The spacer separating these two elements is of conserved length across humans and primates. Productive initiation requires TFAM to engage DNA at a defined distance and rotational orientation relative to the POLRMT/TFB2M recruitment site, since TFAM bends the template and positions POLRMT for promoter melting. The single-nucleotide insertions identified in 9 families, m.16036G>GA and m.16039G>GC, occur within the spacer and alter both the linear distance and the relative positioning of these elements and are therefore expected to reconfigure the pre-initiation complex and modulate the efficiency of light-strand transcription.

**Figure 2.**
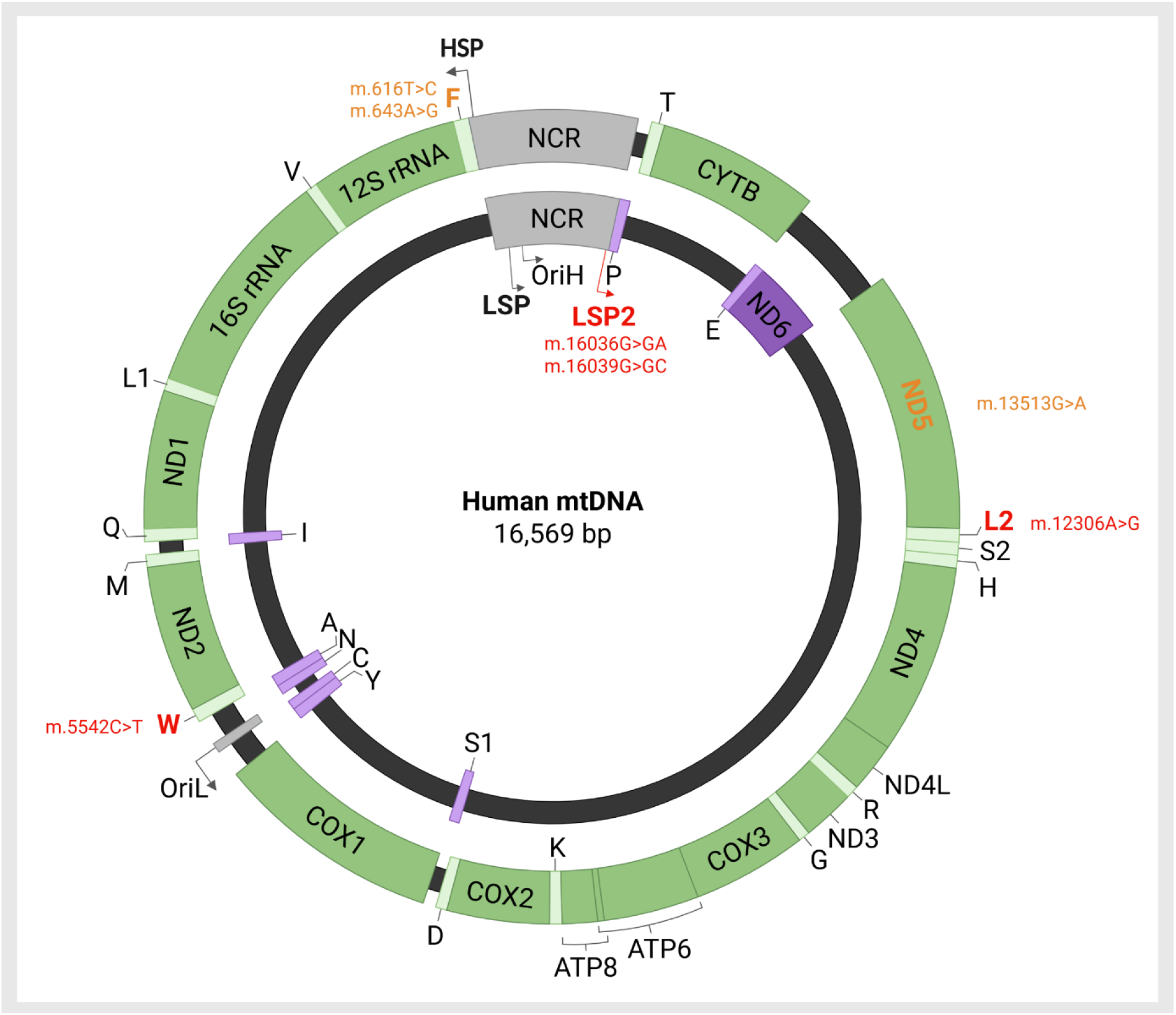
Mitochondrial genome organization and regulation. The double-stranded mitochondrial genome is transcribed from both the heavy (H) and light (L) strands initiating at the corresponding H-strand (HSP) and L-strand (LSP, LSP2) promoters located within the non-coding region (NCR), producing polycistronic transcripts. The NCR region also contains the origin of H-strand replication (OriH). Genes transcribed from the heavy-strand promoter (HSP) and their corresponding gene products are placed in the outer circle and shaded in green. Genes transcribed from the light-strand promoter (LSP or LSP2) and their corresponding gene products are placed in the inner circle and shaded in purple. 22 mitochondrial tRNAs, labeled with their respective amino acid codes, are shown in light purple or light green. Variants previously reported in association with hereditary kidney disease are shown in orange. Newly identified kidney disease-associated variants are shown in red. NCR, non-coding region; CYTB, cytochrome b; ND, NADH dehydrogenase subunits; COX, cytochrome c oxidase subunits; OriH, origin of H-strand replication; Oril, origin of L-strand replication; rRNA, ribosomal RNA. Created in BioRender.com

**Figure 3.**
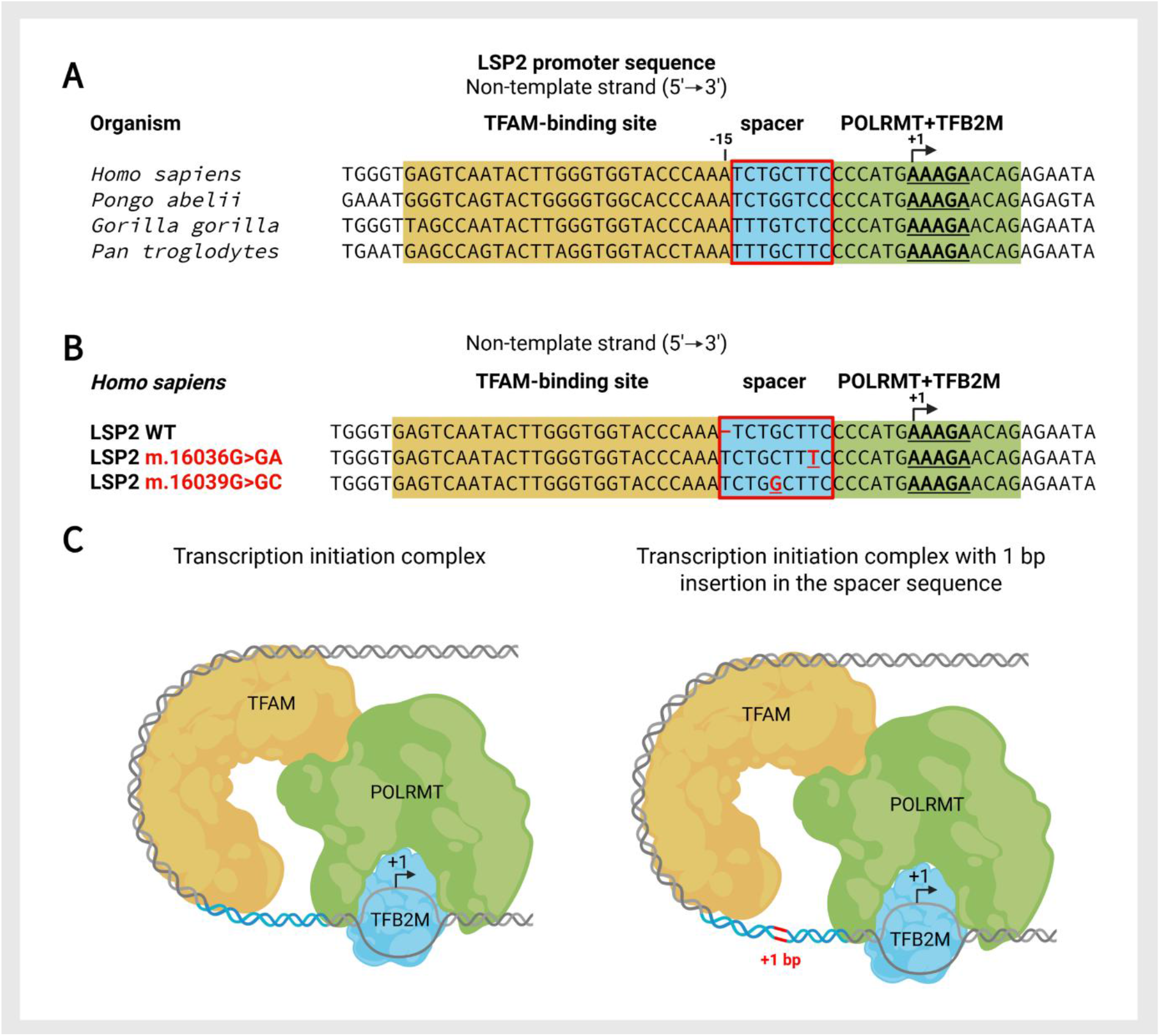
Predicted structural effects of disease-associated single-nucleotide insertions in the mitochondrial light strand promoter 2 (LSP2). **(A)** Sequence alignment of LSP2 (non-template strand) among humans and primates. Red frame highlights the conserved length of the spacer between the TFAM and POLRMT/TFB2M binding sites. The transcription start site (+1) and transcriptional direction (arrow) are indicated. **(B)** Sequence alignment (non-template strand) of the human wild-type LSP2 and the identified variants m.16036G>GA and m.16039G>GC. These insertions increase the distance between the TFAM and POLRMT/TFB2M binding sites. **(C)** Simplified schematic illustrating the position of LSP2 mutation in context of the mitochondrial transcription initiation complex. Created in BioRender.com

#### Disease-Associated mt-tRNA Variants

Transfer RNA (tRNA) is the adaptor molecule that translates the genetic information encoded in mRNA into the amino acid sequence of a protein. The canonical mitochondrial tRNA structure comprises a 7-bp acceptor stem responsible for amino acid attachment, a 5-bp anticodon stem that supports the decoding region, and a 7-nucleotide anticodon loop that mediates codon recognition during protein synthesis.^31^ Furthermore, a conserved non-canonical interaction between positions 32 and 38 stabilizes the anticodon loop through base pairing between its first and last nucleotides. Disruption of this 32–38 interaction can alter the conformation of the anticodon loop, impair codon recognition, and reduce the efficiency of mitochondrial translation. In wild-type mt-tRNA^Phe^, pseudouridine (Ψ) is at position 39.^17^ The variant m.616T>C is predicted to prevent this modification and disrupts base-pairing within the anticodon stem mt-tRNA^Phe^ (**Figure 4A**). The variant m.643A>G disrupts the base-pairing within the acceptor stem of mt-tRNA^Phe^ (**Figure 4A**). The variant m.5542C>T affects position 32 and may therefore perturb the conserved 32-38 interaction stabilizing the anticodon loop of mt-tRNA^Trp^ (**Figure 4B**). Finally, the variant m.12306A>G disrupts base pairing within the anticodon stem of mt-tRNA^Leu(CUN)^ (**Figure 4C**). Collectively, all these disease-associated mt-tRNA variants are predicted to destabilize canonical tRNA secondary structure elements – either the acceptor or anticodon stem, or the conserved anticodon-loop tertiary interaction – representing well-established pathogenic mechanisms in mt-tRNA-related disease.

**Figure 4.**
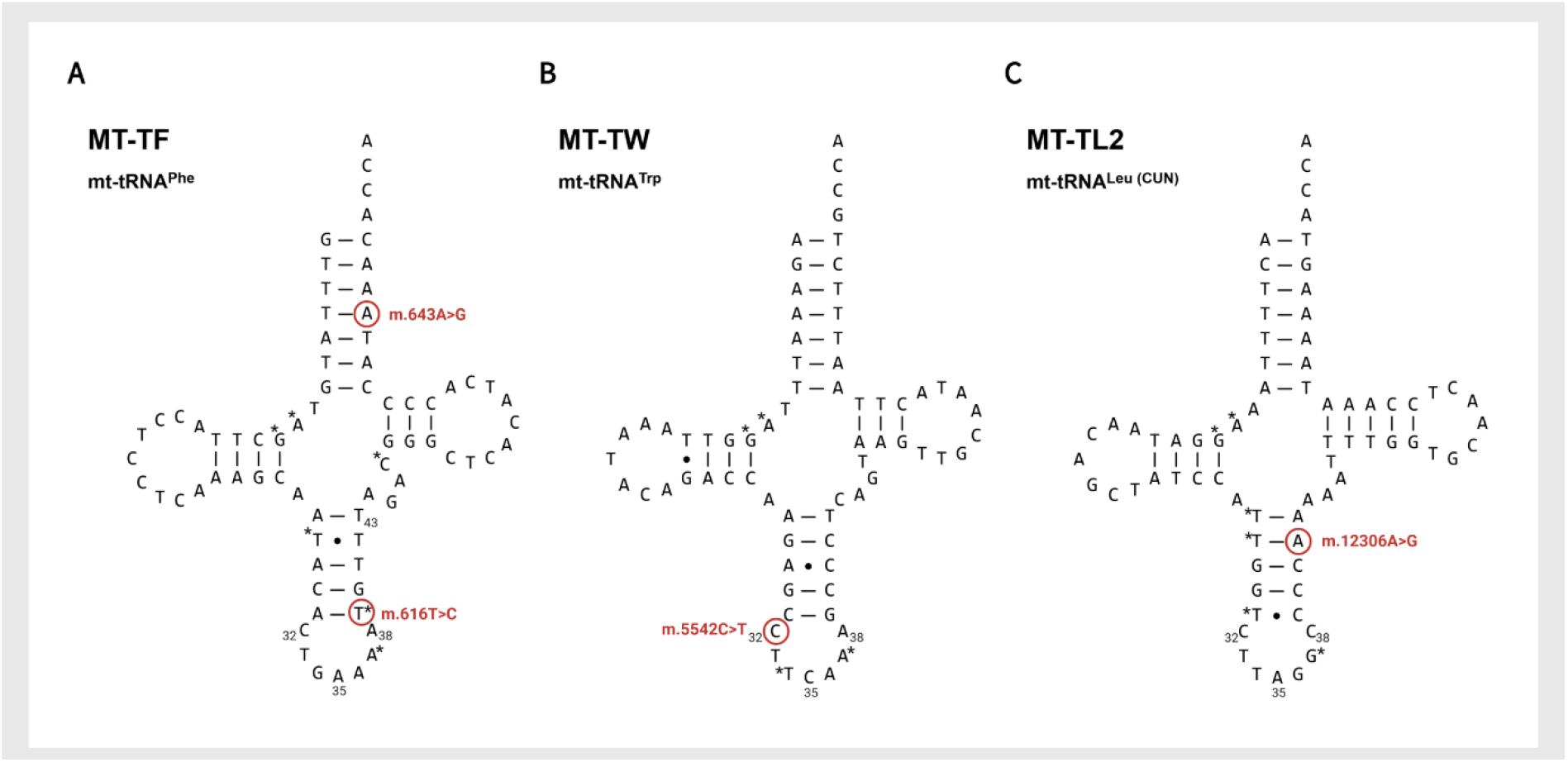
Structural localization of disease-associated variants in mitochondrial tRNAs. **(A)** Predicted secondary structure of mt-tRNA^Phe^ with the m.616T>C and m.643A>G variants located in the anticodon and acceptor stems, respectively. **(B)** Predicted secondary structure of mt-tRNA^Trp^ disrupting the conserved 32-38 interaction that stabilizes the anticodon loop. **(C)** Predicted secondary structure of mt-tRNA^Leu(CUN)^ showing the m.12306A>G variant disrupting base pairing within the anticodon stem. Modified bases are indicated by asterisks. Structures were adapted from Lake et al.^16^; modification sites from Suzuki et al.^17^

### Effects of LSP2 single-nucleotide insertions on i*n vitro* transcription

**Figure 5** shows that spacer length modulates LSP2 promoter activity *in vitro* in a recombinant human mitochondrial transcription system using purified POLRMT, TFAM, and TFB2M.^11^ The single-nucleotide insertions identified in affected families increased LSP2 transcription relative to wild-type, with HSP included as a reference (**Figure 5A**). *In vitro* transcription from dual-promoter templates, in which LSP2 and HSP are arranged in opposite orientations on the same DNA molecule, allowing direct quantitative comparison of HSP- and LSP2-derived transcription within a single reaction, confirmed a four-fold increase in LSP2-derived transcripts for both disease-associated variants compared with wild type sequence, whereas HSP-derived transcripts remained unaffected, with no significant difference between the disease-associated variants (**Figure 5B, C**). The two single-nucleotide insertions identified differ in nucleotide identity but both extend the spacer by one base pair, indicating that promoter activity is determined by spacer length rather than by the identity of the inserted nucleotide. A systematic spacer-length analysis using the same dual-promoter template supported this interpretation: maximal LSP2 activity was observed with insertions of one to two base pairs relative to the native LSP2 spacer, whereas the native HSP spacer length appeared to be near-optimal. Together, these findings demonstrated that both patient-derived single-nucleotide insertions increase LSP2 promoter activity *in vitro* (**Figure 5D**).

**Figure 5.**
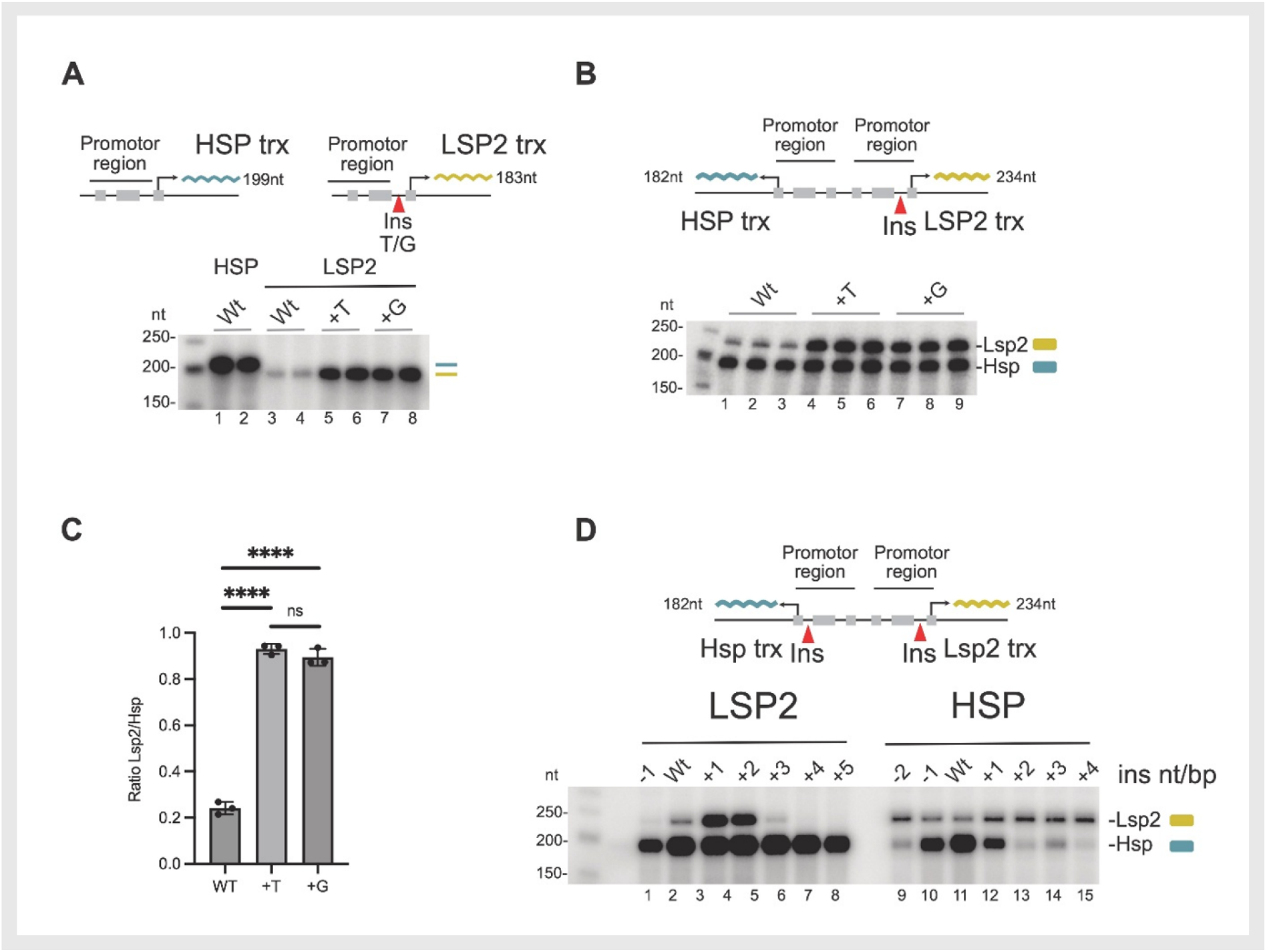
Spacer length modulates LSP2 promoter activity *in vitro*. **(A)** In vitro transcription from single-promoter templates containing either HSP or LSP2, generating transcripts of 199 nt (blue) and 183 nt (yellow), respectively. Single-nucleotide insertions identified in affected families within the LSP2 spacer region are annotated relative to the reference H-strand sequence (NC_012920.1). The m.16036G>GA and m.16039G>GC variants (designated +T and +G, respectively) increased LSP2-derived transcript levels compared with wild type (Wt). **(B)** In vitro transcription from dual-promoter templates in which LSP2 and HSP are arranged in opposite orientations on the same DNA molecule, generating HSP- and LSP2-derived transcripts of 182 nt and 234 nt, respectively. Both +T and +G insertions within the LSP2 spacer increased LSP2-derived transcripts compared with Wt, whereas HSP-derived transcripts were unaffected. **(C)** Quantification of transcription from dual-promoter templates shown in panel B. LSP2 transcription was normalized to HSP within the same reaction. Single-nucleotide insertions in the LSP2 spacer increased promoter activity ∼4-fold, with no detectable difference between +T and +G insertions. Data are presented as mean ± SEM (n = 3). ****p < 0.0001 (one-way ANOVA with Tukey’s multiple-comparisons test). **(D)** Analysis of spacer-length variants using dual-promoter templates. Deletion of a single nucleotide (-1), Wt spacer length, and gradual increases in spacing (+1 to +5 nt) between promoter elements and the TFAM-binding site modulated transcriptional output. The canonical (Wt) LSP2 spacer length was suboptimal, with increased activity observed after insertion of +1 or +2 nt, whereas maximal activity was detected at the canonical (Wt) HSP spacer length.

### Effects of identified mitochondrial DNA variants on mitochondrial proteosynthesis and abundance of respiratory chain complexes

Mitochondrial translation was assessed in patient-derived skin fibroblasts by incorporation of [³⁵S]-methionine and cysteine into newly synthesized mtDNA-encoded proteins. Cell lines harboring either of the LSP2 single-nucleotide insertions showed similarly reduced mitochondrial protein synthesis (**Figure 6A**), whereas the decrease was less pronounced in fibroblasts carrying the *MT-TF* m.616T>C or *MT-TW* m.5542C>T variant (**Figure 6B**). This was confirmed by signal quantification, which revealed a significant reduction in the levels of complex IV subunits, particularly MT-CO1 and MT-CO2, as well as the complex III subunit MT-CYB, an effect that was less evident in cells carrying the *MT-TW* m.5542C>T variant (**Figure 6C**). Although pulse labeling demonstrated impaired mitochondrial translation, signals after a 20-hour chase were comparable between controls and all patient cell lines. When expressed as the pulse/chase signal ratio, patient fibroblasts — most prominently those carrying the LSP2 variants — showed reduced turnover of mitochondrial proteins, possibly reflecting a compensatory mechanism (**Figure 6D**).

**Figure 6.**
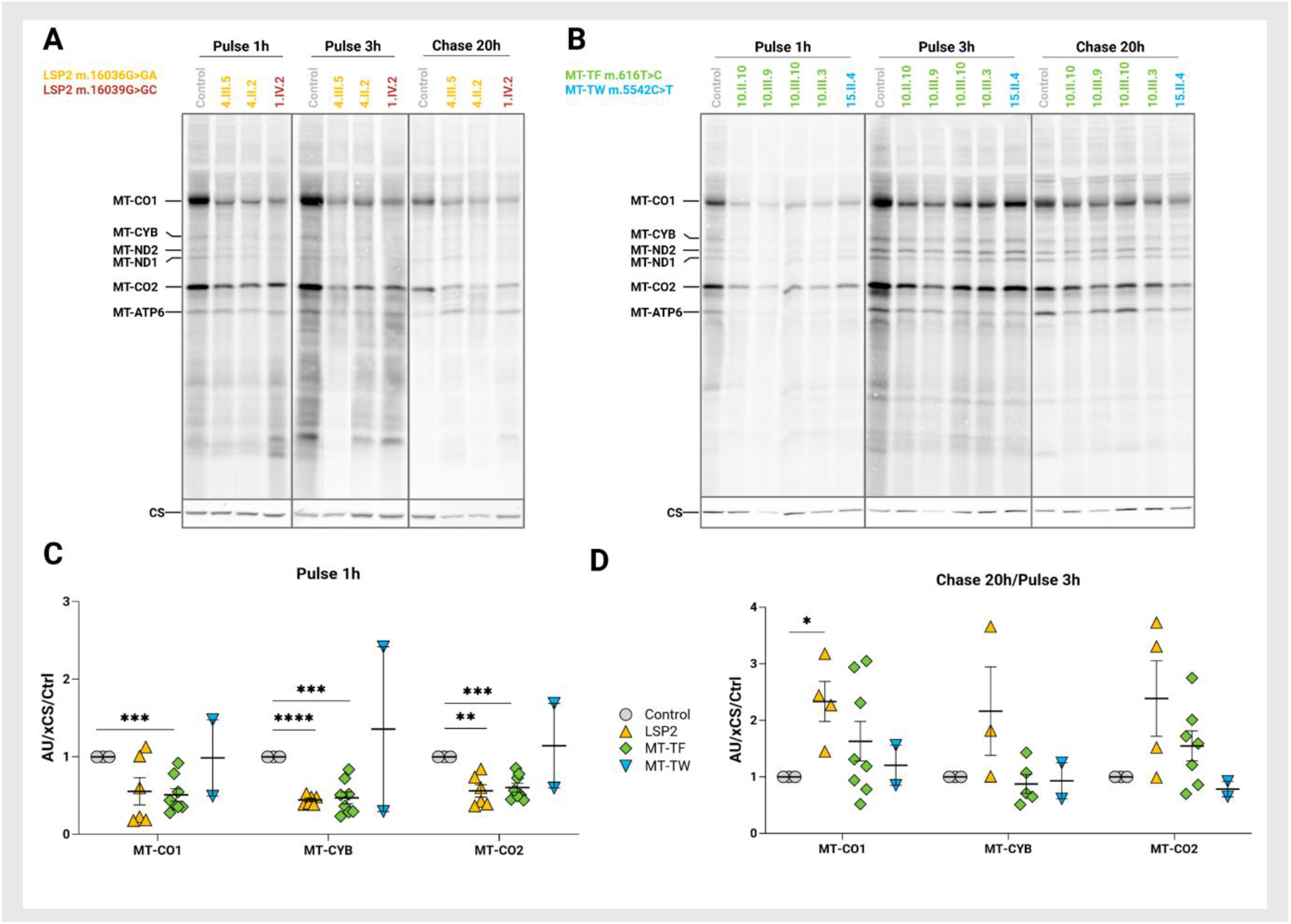
Mitochondrial proteosynthesis in LSP2-, *MT-TF*-, and *MT-TW*-derived patient skin fibroblasts. **(A)** Representative autoradiographs of mtDNA-encoded OXPHOS subunits labeled with [^35^S]-methionine and cysteine for 1 or 3 h in fibroblasts carrying either of the two LSP2 single-nucleotide insertions, and after a 20 h chase, analyzed by SDS-PAGE. (B) Representative autoradiographs of [^35^S]-labeled mtDNA-encoded OXPHOS subunits in fibroblasts carrying the *MT-TF* m.616T>C or *MT-TW* m.5542C>T variant, analyzed in the same manner. (C) Quantification of 1 h pulse [^35^S] in vivo labeling signals for representative complex IV subunits (MT-CO1 and MT-CO2) and the complex Ill subunit MT-CYB, normalized to citrate synthase (CS). (D) Quantification of the 20 h chase-to-3 h pulse ratio of [^35^S] in vivo labeling signals for representative complex IV subunits (MT-CO1 and MT-CO2) and the complex Ill subunit MT-CYB. Signal of controls was normalized to 1. Data for patient fibroblasts are presented as mean ± SEM. *p < 0.05, **p < 0.01, ***p < 0.001, ****p < 0.0001 (one-sample t-test against the controls mean of 1).

Consequently, blue native gel electrophoresis (BNE) demonstrated a selective decrease in complexes I and IV abundance in fibroblasts carrying both LSP2 variants. In contrast, this reduction was less pronounced or not observed in fibroblasts harboring either the *MT-TF* m.616T>C or the *MT-TW* m.5542C>T variant individually (**Figure 7A, B**). Untargeted proteomic analysis confirmed a selective reduction in mtDNA-encoded proteins and in the abundance of complexes I and IV, as inferred from the reduced abundance of their constituent subunits, in fibroblasts carrying both LSP2 variants, while the levels of housekeeping nuclear-encoded proteins remained unchanged (**Figure 7C, D; Supplementary Figure S2**). A similar pattern was observed in fibroblasts harboring the *MT-TW* m.5542C>T variant alone. In contrast, the *MT-TF* m.616T>C variant did not affect mtDNA-encoded protein levels but was associated with a specific reduction in complex IV abundance, based on the decreased levels of its component proteins (**Figure 7C, D; Supplementary Figure S3**).

**Figure 7.**
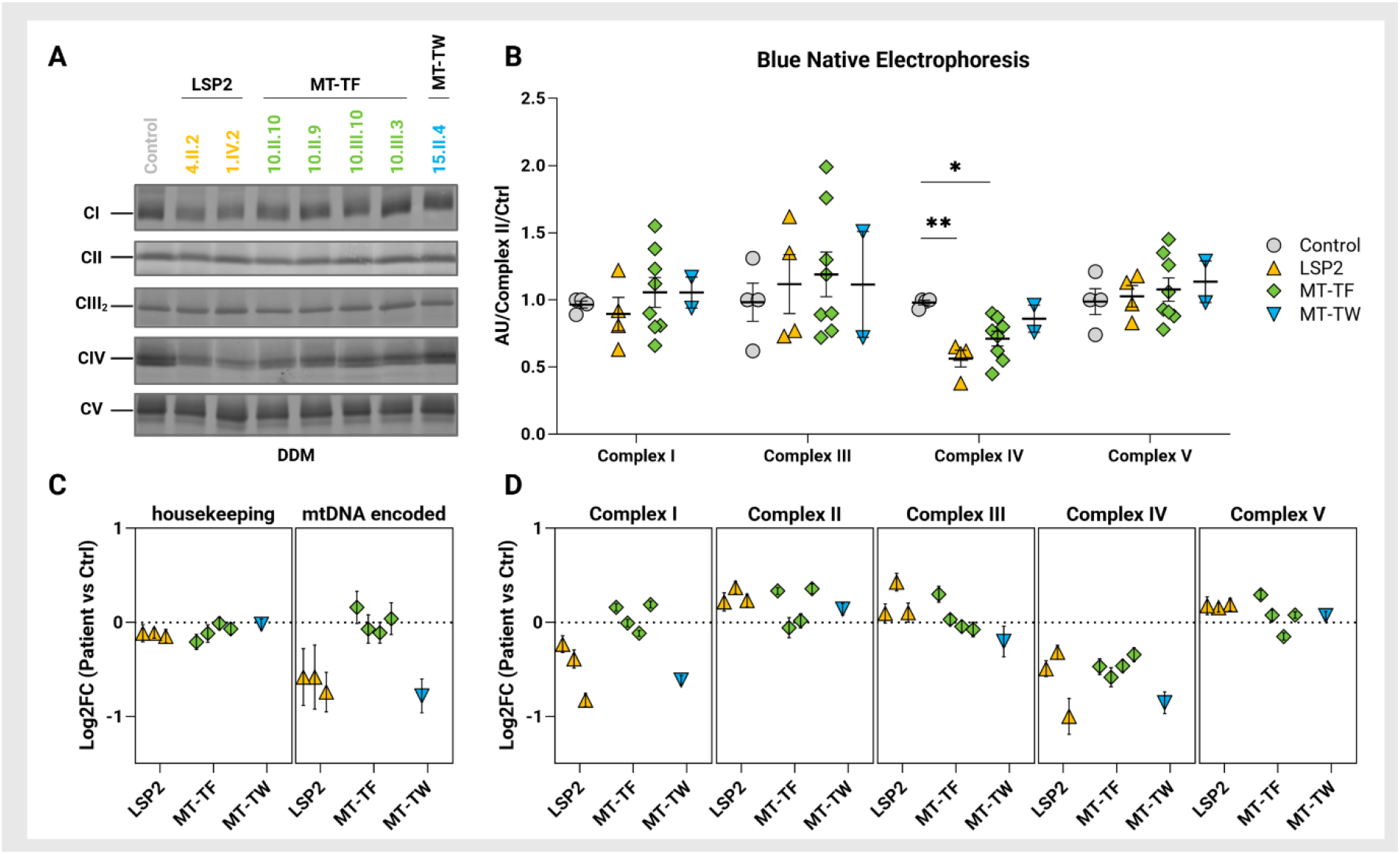
Respiratory chain complex abundance in LSP2-, *MT-TF*-, and *MT-TW*-derived patient skin fibroblasts. **(A)** Representative blue native electrophoresis (BNE) gel of the whole cells solubilized by n-dodecyl β-D-maltoside (DDM). **(B)** Quantification of BNE respiratory chain complex abundance, normalized to complex Il. Statistical analysis was performed for each complex using one way ANOVA *p < 0.05, **p < 0.01. **(C)** Mean log_2_, fold changes ± SEM of all detected housekeeping and mtDNA-encoded proteins determined by untargeted proteomic analysis. **(D)** Mean log_2_, fold changes ± SEM of all detected structural subunits within individual OXPHOS complexes determined by untargeted proteomic analysis.

### Effects of identified mitochondrial DNA variants on respiratory chain activity and CoQ10 content

Consistent with impaired mitochondrial protein synthesis and respiratory chain complex assembly, oxygen consumption was reduced in all patient-derived fibroblasts, with the exception of the cell line from individual 1.IV.2 carrying the m.16039G>GC LSP2 variant, which exhibited marked variability. Reduced respiration was observed both under coupled conditions (OXPHOS; **Figure 8A**), reflecting ATP-generating respiratory capacity, and at maximal respiratory capacity (ETC; **Figure 8B**), reflecting the maximal electron transport capacity of the respiratory chain. Complex IV activity measured by high-resolution respirometry (COX; **Figure 8C**) showed a similar pattern of reduction, further supporting impaired respiratory chain function. In contrast, the uncoupling control ratio (UCR; **Figure 8D**) remained unchanged in fibroblasts carrying LSP2 variants, indicating that ATP synthase activity was not rate-limiting. In comparison, fibroblasts carrying the *MT-TF* and *MT-TW* variants showed a partial reduction in UCR, suggesting a limitation at the level of respiratory chain rather than at the components of the phosphorylation system. Nevertheless, the overall decrease in OXPHOS, ETC, and COX activities suggested that the primary respiratory defect resulted from a combined respiratory chain deficiency, consistent with impaired mitochondrial protein synthesis and reduced abundance of individual OXPHOS complexes. Along with impaired mitochondrial translation and respiratory chain function, LSP2-, *MT-TF*-, and *MT-TW*-derived fibroblasts exhibited significantly reduced CoQ10 levels compared with controls. This decrease persisted after normalization to citrate synthase activity, indicating that it was not attributable to differences in mitochondrial content (**Figure 8E, F**).

**Figure 8.**
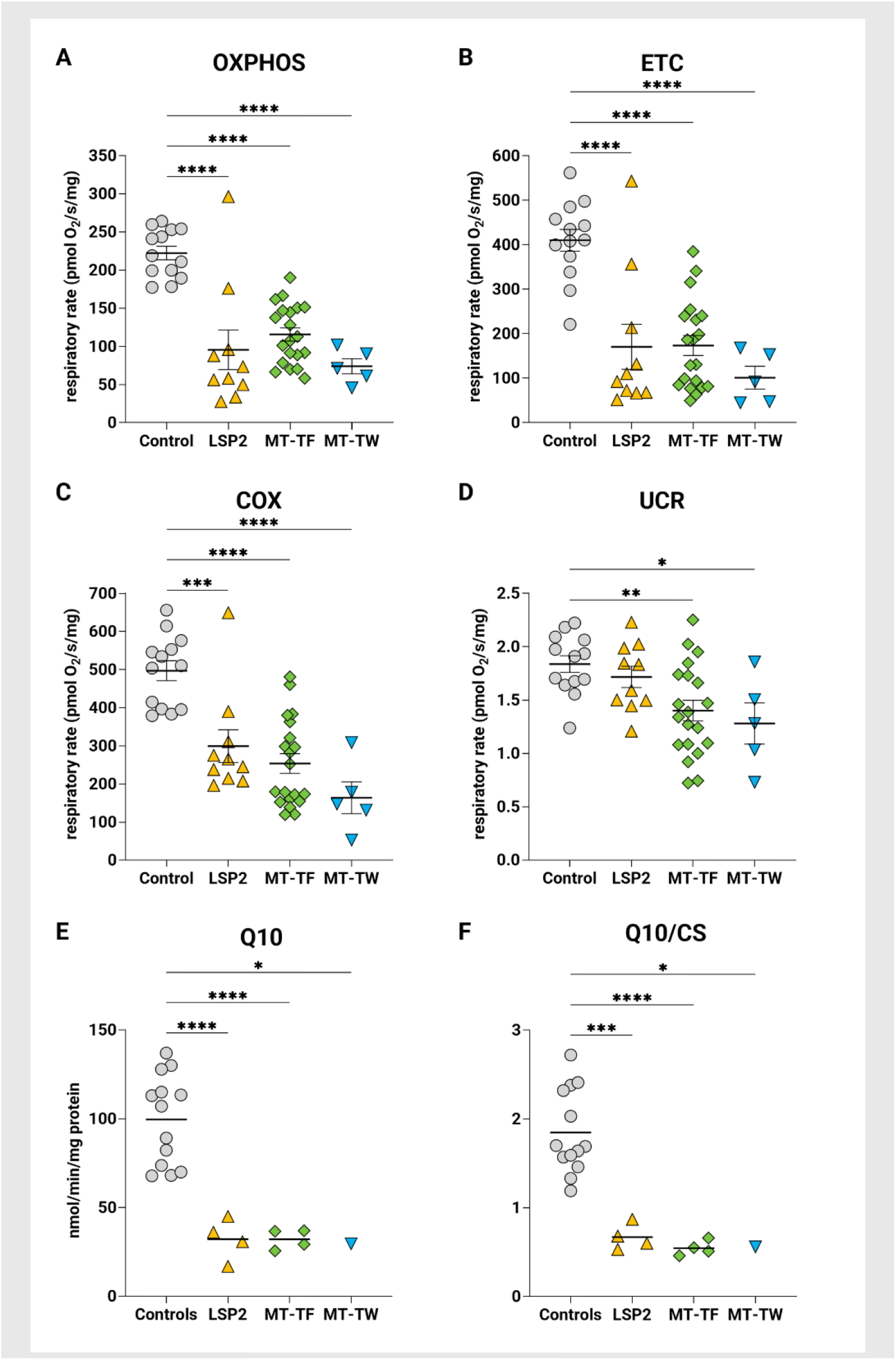
Respiratory chain function and CoQ10 levels in LSP2-, *MT-TF*-, and *MT-TW*-derived patient skin fibroblasts. **(A)** Oxygen consumption under coupled conditions (OXPHOS), reflecting ATP-generating respiratory capacity. **(B)** Maximal respiratory capacity (ETC), reflecting the maximal electron transport capacity of the respiratory chain. **(C)** Complex IV activity (COX) measured by high-resolution respirometry using artificial substrates. (D) Uncoupling control ratio (UCR), representing the ratio of maximal respiratory capacity to OXPHOS respiration. **(E)** CoQ10 levels. **(F)** CoQ10 levels normalized to citrate synthase (CS) activity. Each dot represents the mean of two technical measurements, and horizontal lines indicate group means ± SEM. Statistical analysis was performed using one-way ANOVA with multiple comparison test. *p < 0.05, **p < 0.01, ***p < 0.001, ****p < 0.0001.

## Discussion

In this study, we evaluated mitochondrial DNA variation in 33 families with suspected ADTKD from the WF-CUNI-RIKD registry.^12, 13^ These families had previously tested negative in genetic analyses focused on autosomal dominant inheritance but, upon clinical re-evaluation, showed features consistent with maternal inheritance.

Eighteen (55%) families were found to carry seven types of mitochondrial disease-associated variants. These include recurrent single-nucleotide insertions in the second light-strand promoter (LSP2)^11^ observed in nine families, a *MT-TW* variant in two families, and a variant in *MT-TL2* in one family, along with three disease-associated variant types in *MT-TF* and *MT-ND5*.^6, 9, 10^ In 16 families, disease-associated variants occurred on distinct haplotypes, consistent with independent mutational events, whereas two families with the m.5542C>T variant in *MT-TW* shared haplogroup U2e2a1, suggesting distant shared ancestry. All disease-associated variant types were present in blood at near-complete or complete homoplasmy, except for the *MT-ND5* variant, which was detected at a heteroplasmy level of 5–10%. Maternal inheritance typical of mitochondrial disease was supported in these families, as only 1 of 17 children of genetically affected fathers had below-normal kidney function, whereas 54 of 60 children of affected mothers were clinically affected (p = 1.23 × 10⁻^11^). The pathogenicity of the individual disease-associated variant types was further supported by their predicted deleterious structural effects and by functional evidence demonstrating impaired mitochondrial transcription, reduced mitochondrial protein synthesis, respiratory chain deficiency, and CoQ10 depletion.

In total, 54/60 genetically affected individuals and 55/59 obligate at-risk carriers were clinically affected. Eight of the remaining 10 individuals were between ages 20-45 and may not have yet experienced kidney function decline. Clinical status was unknown in an additional 66 obligate at-risk carriers at the time of evaluation. Affected individuals all suffered from chronic tubulointerstitial kidney disease, with the occasional presence of gout and other clinical manifestations of mitochondrial disease, including short stature, seizures, myopathy and hearing loss. However, these non-renal presentations were not uniform within the families, but rather case-specific. The rate of CKD progression appeared to vary both between and within families. Kidney histopathology was nonspecific and non-diagnostic.

Our findings have several clinical implications. They expand the spectrum of genetic causes underlying tubulointerstitial kidney disease and support the use of the term MITKD^8^ for mitochondrial forms of the condition. They also highlight the need for a standardized nomenclature, analogous to that recommended by KDIGO for ADTKD,^32^ with gene-specific designations (e.g. MITKD-LSP2 and related entities). The observed disease-associated variant spectrum, arising from independent mutational events, together with extensive pedigrees containing multiple clinically affected individuals, is characteristic for mitochondrial diseases and maternal inheritance. These findings suggest that mitochondrial disease may be a relatively frequent cause of TKD and should be considered in the differential diagnosis of cases with unresolved genetic etiology, particularly when maternal inheritance is suspected. In light of these findings, we suggest the differential diagnostic decision tree outlined in **Figure 9**. While maternal inheritance of TKD alone should be an indication for evaluating mitochondrial causes, other manifestations of mitochondrial disease may also be present, including hearing deficit, optic nerve abnormalities, lactic acidosis, diabetes, and seizures, which are primarily associated with more severe mitochondrial dysfunction.

**Figure 9.**
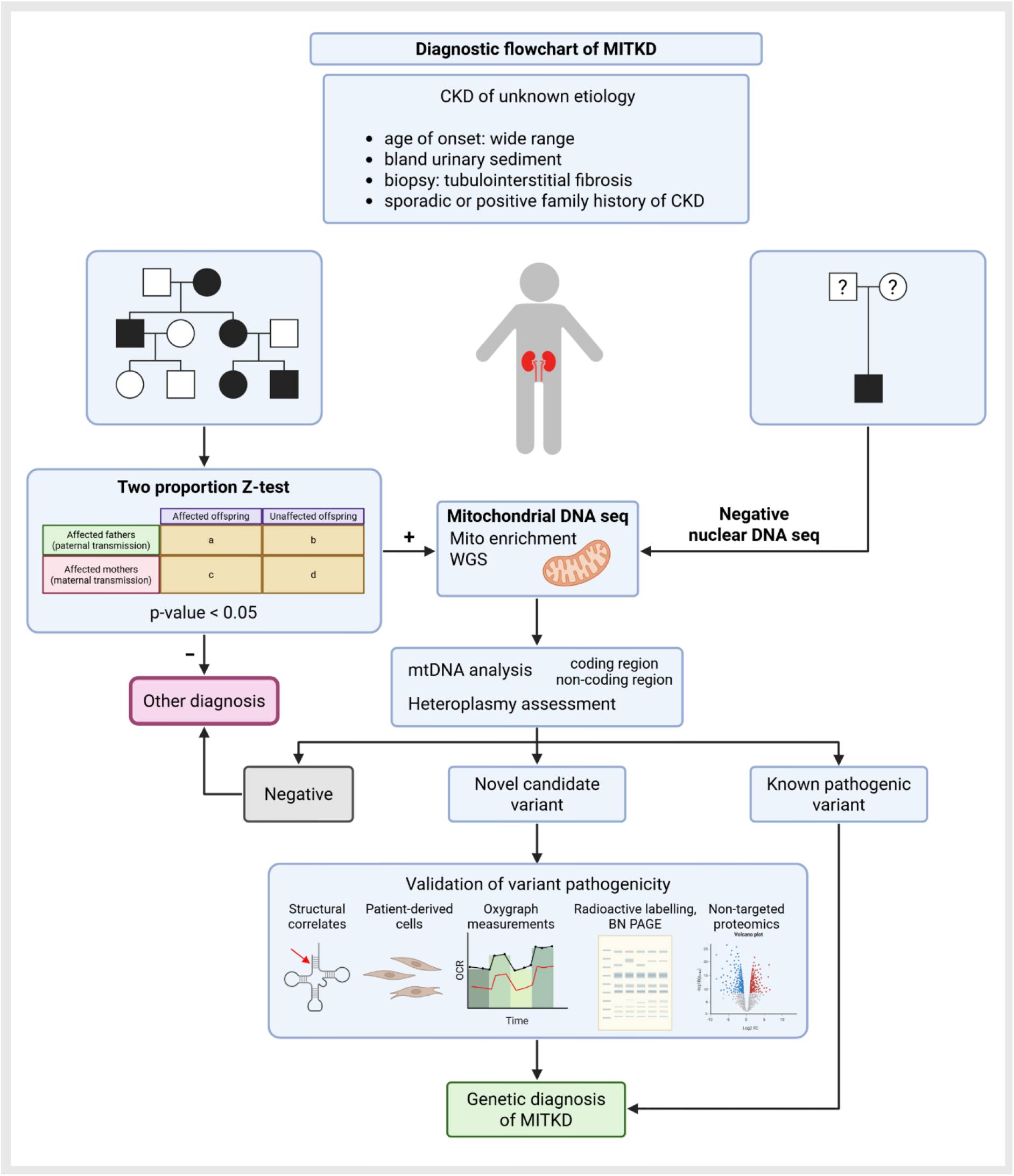
Diagnostic flowchart of MITKD. Mitochondrial inheritance should be considered in both familial and sporadic cases of isolated tubulointerstitial kidney disease of unknown etiology, particularly when routine genetic testing of nuclear DNA is unrevealing. The likelihood of mitochondrial inheritance can be evaluated by comparing the frequency of impaired kidney function among offspring of clinically affected mothers and fathers using two proportion Z-test, with a significant excess among maternal offspring supporting mitochondrial transmission. Mitochondrial DNA sequence and heteroplasmy levels can be assessed through variety of sequencing approaches including targeted and/ or untargeted protocols. Negative results indicate non-mitochondrial inheritance. Novel candidate variants need to be functionally characterized. Created in BioRender.com

One of the key findings in this investigation was the identification of two types of recurrent homoplasmic single-nucleotide insertions in the second light-strand promoter (LSP2), observed in nine families. One of the variants, m.16039G>GC, was previously reported in a single MITKD family in combination with a non-coding homoplasmic m.547A>T variant that had been considered pathogenic based on its localization within the heavy strand promoter (HSP) and functional studies showing reduced expression of heavy strand tRNAs relative to light strand tRNAs, as well as decreased mitochondrial protein synthesis and respiratory chain activity. The pathogenic potential of the m.16039G>GC insertion was likely underappreciated in that study, because it resided within a hypervariable region of the mitochondrial control region downstream of the canonical light-strand promoter, an interval not previously recognized as functionally important.^8^ However, our data question a pathogenic role for the m.547A>G substitution while providing strong genetic and functional support for the biological and clinical relevance of LSP2, based on the exclusive presence of recurrent disease-associated single-nucleotide insertions within this region. These insertions are predicted to alter the LSP2 architecture, resulting in an approximately 4-fold increase in mitochondrial light strand transcription *in vitro*. *In vivo* these variants result in decreased mitochondrial protein synthesis and respiratory chain defect, suggesting a global disruption of mitochondrial transcription and translation through a yet unknown mechanism, that needs to be addressed in future work.

Interestingly, in this study a shift towards homoplasmy was observed for all LSP2 and mt-tRNA variant types, but not for the *MT-ND5* variant affecting the NADH-ubiquinone oxidoreductase chain 5 protein (ND5), a structural subunit of Complex I. Similar shifts to near- or complete homoplasmy have also been reported for mt-tRNA variants in other MITKD families,^8, 9, 33–35^ although mechanisms underlying this dynamic remain poorly understood. To this end, a recent study on three gene-edited mouse models harboring distinct pathogenic mt-tRNA variants demonstrated age-dependent shifts towards homoplasmy specifically in kidney, accompanied by renal dysfunction. In contrast, pathogenic variant load remained stable in brain, muscle, heart and testis and even decreased in blood, spleen and colon. Consistent with these tissue-specific effects, no differences in fertility were observed in either female or male mice. However, pathogenic variant load in offspring increased progressively across generations, in proportion to the maternal pathogenic variant load.^36^ Although the underlying mechanism remains unclear, our findings support the concept that certain pathogenic regulatory and mt-tRNA variants promote gradual, tissue-specific remodeling of the mitochondrial genome also in human. In the kidney, a long-lived, energy-demanding organ with limited cellular turnover, progressive enrichment of dysfunctional mitochondrial genomes may ultimately exceed a functional threshold and result in age-dependent kidney dysfunction. Consistent with mouse data^36^ this process may be further amplified across generations by maternal inheritance and the mitochondrial bottleneck, resulting in increasing variant load and earlier or more severe renal disease in successive generations.

In summary, we show that specific variants in LSP2 and mt-tRNA cause inherited tubulointerstitial kidney disease, particularly in cases with later onset and absence of extrarenal manifestations. These variants are characterized by near-complete or complete homoplasmy in blood, distinguishing them from other mitochondrial disease-associated variants that are pathogenic at low heteroplasmy levels and typically present with early-onset, multisystem involvement. Notably, such variants may accumulate preferentially in the kidney over time and can manifest as apparently sporadic late-onset CKD, as demonstrated in mouse models.^36^ These findings support the inclusion of the mitochondrial genome in inherited kidney disease gene panels and highlight the need for careful evaluation of mitochondrial variants in both familial and sporadic cases of isolated tubulointerstitial kidney disease of unknown etiology, as the clinical presentation may be nonspecific, mitochondrial inheritance may be unrecognized, and penetrance may be incomplete.

## Data Availability

All data produced in the present study are available upon reasonable request to the authors

## Acknowledgements

We would like to thank the Proteomics Service Laboratory at the Institute of Physiology (supported by RVO, ID 67985823) and the Institute of Molecular Genetics (supported by RVO, ID 68378050) of the Czech Academy of Sciences for processing of the proteomics data and to ERDF JAC ACGT2 project (CZ.02.01.01/00/23_020/0008555) from the Ministry of Education, Youth and Sports of the Czech Republic for providing allele frequencies from Czech control genomes.

We sincerely thank the many participating individuals and their families, together with their primary physicians and colleagues, whose generous contributions of clinical data and biological specimens made this work possible. We also thank the following physicians for referring families to the Wake Forest Registry: Louise Amlie-Wolf (Nemours Children’s Hospital, Wilmington, DE, USA), James Bordeau (Nephrology Specialists of Oklahoma, Tulsa, OK, USA), Hui Jen Ding (Kuala Lumpur Hospital, Jalan Pahang, Kuala Lumpur, Malaysia), Balram Gangaram (University of California, San Francisco, CA, USA), Edward Hovick (Kidney Care Specialists, LLC, Broomall, PA, USA), Fred Lui (Hill Physicians Medical Group, Burlingame, CA, USA), Laura Nishi (Northwestern Medical Group, Chicago, IL), Rohini Prashar (Henry Ford Health, Detroit, MI, USA), Rose Shim (Ohio State University Wexner Medical Center, Columbus, OH, USA), and Andrew Talbot (Melbourne Kidney Specialists, Richmond, Victoria, Australia). Their commitment has been invaluable and continues to advance our understanding of hereditary TKD.

## Author Contribution Statement

KS, KK, MŽ, AJB and SK conceived the concept, designed and coordinated the study and wrote the original manuscript draft. DM, TK, HHart, KH, VS, VB, HT, LP, JS, HŠ, HHans, KT, PP, VK, MV, TM, OP, CMG, and MF were responsible for acquisition and analysis of data. KK, AT, LM, AS, NW, ES and AJB organized, curated and maintained the WF-CUNI-RIKD registry. KK, MM, JZ, KS, AS and AJB collected and curated clinical materials and associated patient information for research and registry purposes. All authors reviewed the results and approved the final version of the manuscript. The results presented in this article have not been published previously in whole or in part, except in abstract form.

## Data sharing statement

This is a clinical study, and individual-level clinical and genetic data cannot be shared to protect participant privacy. The mass spectrometry proteomics data have been deposited to the ProteomeXchange Consortium via the PRIDE partner repository with the dataset identifier PXD079905.

## Declaration of Generative AI and AI-assisted technologies in the manuscript preparation process

The AI tools were used solely to improve the grammar of the text.

## References

1. Elhassan EAE, Cormican S, Osman SM, et al. Characterization of Monogenic Kidney Disease in Older Patients With CKD. Kidney Int Rep 2025; 10: 2140–2152.

2. Popp B, Ekici AB, Knaup KX, et al. Prevalence of hereditary tubulointerstitial kidney diseases in the German Chronic Kidney Disease study. Eur J Hum Genet 2022; 30: 1413–1422.

3. Groopman EE, Marasa M, Cameron-Christie S, et al. Diagnostic Utility of Exome Sequencing for Kidney Disease. N Engl J Med 2019; 380: 142–151.

4. Bleyer AJ, Kidd KO, Zivna M, et al. Autosomal Dominant Tubulointerstitial Kidney Disease: A Review. Am J Kidney Dis 2025; 86: 677–689.

5. Tzen CY, Tsai JD, Wu TY, et al. Tubulointerstitial nephritis associated with a novel mitochondrial point mutation. Kidney Int 2001; 59: 846–854.

6. Zsurka G, Hampel KG, Nelson I, et al. Severe epilepsy as the major symptom of new mutations in the mitochondrial tRNA(Phe) gene. Neurology 2010; 74: 507–512.

7. D’Aco KE, Manno M, Clarke C, et al. Mitochondrial tRNA(Phe) mutation as a cause of end-stage renal disease in childhood. Pediatr Nephrol 2013; 28: 515–519.

8. Connor TM, Hoer S, Mallett A, et al. Mutations in mitochondrial DNA causing tubulointerstitial kidney disease. PLoS Genet 2017; 13: e1006620.

9. Viering D, Schlingmann KP, Hureaux M, et al. Gitelman-Like Syndrome Caused by Pathogenic Variants in mtDNA. J Am Soc Nephrol 2022; 33: 305–325.

10. Bakis H, Trimouille A, Vermorel A, et al. Adult onset tubulo-interstitial nephropathy in MT-ND5-related phenotypes. Clin Genet 2020; 97: 628–633.

11. Tan BG, Mutti CD, Shi Y, et al. The human mitochondrial genome contains a second light strand promoter. Mol Cell 2022; 82: 3646–3660 e3649.

12. Bleyer AJ, Kidd K, Robins V, et al. Outcomes of patient self-referral for the diagnosis of several rare inherited kidney diseases. Genet Med 2020; 22: 142–149.

13. Olinger E, Hofmann P, Kidd K, et al. Clinical and genetic spectra of autosomal dominant tubulointerstitial kidney disease due to mutations in UMOD and MUC1. Kidney Int 2020; 98: 717–731.

14. Astley ME, Chesnaye NC, Hallan S, et al. Age- and sex-specific reference values of estimated glomerular filtration rate for European adults. Kidney Int 2025; 107: 1076–1087.

15. Thompson JD, Gibson TJ, Higgins DG. Multiple sequence alignment using ClustalW and ClustalX. Curr Protoc Bioinformatics 2002; Chapter 2: Unit 2 3.

16. Lake NJ, Zhou L, Xu J, et al. MitoVisualize: a resource for analysis of variants in human mitochondrial RNAs and DNA. Bioinformatics 2022; 38: 2967–2969.

17. Suzuki T, Yashiro Y, Kikuchi I, et al. Complete chemical structures of human mitochondrial tRNAs. Nat Commun 2020; 11: 4269.

18. Posse V, Hoberg E, Dierckx A, et al. The amino terminal extension of mammalian mitochondrial RNA polymerase ensures promoter specific transcription initiation. Nucleic Acids Res 2014; 42: 3638–3647.

19. Posse V, Shahzad S, Falkenberg M, et al. TEFM is a potent stimulator of mitochondrial transcription elongation in vitro. Nucleic Acids Res 2015; 43: 2615–2624.

20. Hartmannova H, Piherova L, Tauchmannova K, et al. Acadian variant of Fanconi syndrome is caused by mitochondrial respiratory chain complex I deficiency due to a non-coding mutation in complex I assembly factor NDUFAF6. Hum Mol Genet 2016; 25: 4062–4079.

21. Cunatova K, Reguera DP, Vrbacky M, et al. Loss of COX4I1 Leads to Combined Respiratory Chain Deficiency and Impaired Mitochondrial Protein Synthesis. Cells 2021; 10.

22. Fernandez-Vizarra E, Zeviani M. Blue-Native Electrophoresis to Study the OXPHOS Complexes. Methods Mol Biol 2021; 2192: 287–311.

23. Cunatova K, Vrbacky M, Knezu M, et al. The cytochrome c oxidase subunit COX6B1 is required for redox-sensitive early assembly and late stabilization of complex IV. J Biol Chem 2026; 302: 111070.

24. Bartosova T, Klempir J, Hansikova H. Coenzyme Q10: A Biomarker in the Differential Diagnosis of Parkinsonian Syndromes. Antioxidants (Basel) 2023; 12.

25. Krizova J, Hulkova M, Capek V, et al. Microarray and qPCR Analysis of Mitochondrial Metabolism Activation during Prenatal and Early Postnatal Development in Rats and Humans with Emphasis on CoQ(10) Biosynthesis. Biology (Basel) 2021; 10.

26. Srere PA. [1] Citrate synthase: [EC 4.1.3.7. Citrate oxaloacetate-lyase (CoA-acetylating)]. Methods in Enzymology, vol. 13. Academic Press, 1969, pp 3–11.

27. Alan L, Opletalova B, Hayat H, et al. Mitochondrial metabolism and hypoxic signaling in differentiated human cardiomyocyte AC16 cell line. Am J Physiol Cell Physiol 2025; 328: C1571–C1585.

28. Tyanova S, Temu T, Sinitcyn P, et al. The Perseus computational platform for comprehensive analysis of (prote)omics data. Nat Methods 2016; 13: 731–740.

29. Tan BG, Gustafsson CM, Falkenberg M. Mechanisms and regulation of human mitochondrial transcription. Nat Rev Mol Cell Biol 2024; 25: 119–132.

30. Clyde D. Promoting a new view of mitochondrial genome regulation. Nat Rev Genet 2022; 23: 648–649.

31. Salinas-Giege T, Giege R, Giege P. tRNA biology in mitochondria. Int J Mol Sci 2015; 16: 4518–4559.

32. Eckardt KU, Alper SL, Antignac C, et al. Autosomal dominant tubulointerstitial kidney disease: diagnosis, classification, and management--A KDIGO consensus report. Kidney Int 2015; 88: 676–683.

33. Lorenz R, Ahting U, Betzler C, et al. Homoplasmy of the Mitochondrial DNA Mutation m.616T>C Leads to Mitochondrial Tubulointerstitial Kidney Disease and Encephalopathia. Nephron 2020; 144: 156–160.

34. Buglioni A, Hasadsri L, Nasr SH, et al. Mitochondriopathy Manifesting as Inherited Tubulointerstitial Nephropathy Without Symptomatic Other Organ Involvement. Kidney Int Rep 2021; 6: 2514–2518.

35. Xu C, Tong L, Rao J, et al. Heteroplasmic and homoplasmic m.616T>C in mitochondria tRNAPhe promote isolated chronic kidney disease and hyperuricemia. JCI Insight 2022; 7.

36. Zhang L, Xu Z, Jing J, et al. Age-dependent accumulation of mitochondrial tRNA mutations in mouse kidneys linked to mitochondrial kidney diseases. Nat Aging 2025; 5: 1317–1339.

37. Wong LC, Chen T, Wang J, et al. Interpretation of mitochondrial tRNA variants. Genet Med 2020; 22: 917–926.

38. Barone V, La Morgia C, Caporali L, et al. Case Report: Optic Atrophy and Nephropathy With m.13513G>A/MT-ND5 mtDNA Pathogenic Variant. Front Genet 2022; 13: 887696.

