## Supplementary Material for "Recurrent Single-Nucleotide Insertions in the Mitochondrial Second Light-Strand Promoter Cause Tubulointerstitial Kidney Disease"

This appendix has been provided by the authors to give readers additional information about their work.

**Table of contents**

**Supplementary Figure S1** *(p.2-3)*

Representative histopathological findings in renal biopsies of patients with MITKD.

**Supplementary Figure S2** *(p.4)*

Non-targeted proteomic analysis of fibroblasts from patients with LSP2 (4.III.5; 4.II.2; 1.IV.2) variants.

**Supplementary Figure S3** *(p.5-6)*

Non-targeted proteomic analysis of fibroblasts from patients with *MT-TF* (10.II.10; 10.III.9; 10.III.10; 10.III.3) and *MT-TW* (15.II.4) variants.

**Supplementary Table S1** *(p.7)*

Comparison of maternal vs. non-maternal clinical data from 224 adults identified by pedigrees.

**Supplementary Methods** *(p.8-14)*

**Supplementary References** *(p.15-16)*

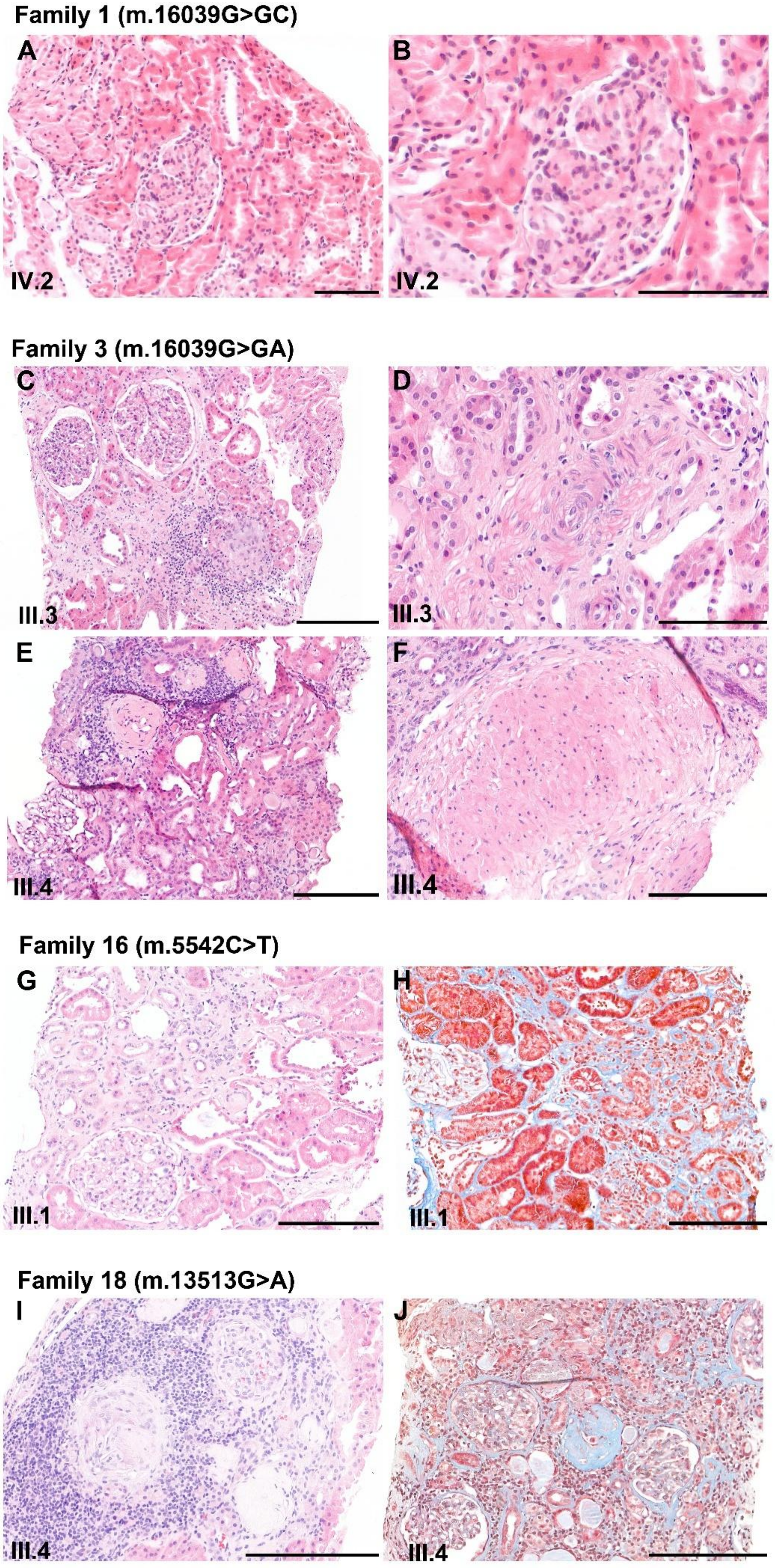
Family 1 (m.16039G>GC)
A
B
IV.2
IV.2
Family 3 (m.16039G>GA)
C
D
III.3
III.3
E
F
III.4
III.4
Family 16 (m.5542C>T)
G
H
III.1
III.1
Family 18 (m.13513G>A)
I
J
III.4
III.4

**Supplementary Figure S1: Representative histopathological findings in renal biopsies of patients with MITKD. (A, B)** Patient IV.2 (family 1), biopsy at 36-40 yrs. Biopsy showed mild mesangial hypercellularity in glomeruli, otherwise insignificant changes. **(C, D)** Patient III.3 (family 3), biopsy at 16-20 yrs. Moderate interstitial fibrosis, tubular atrophy (C) and smooth muscle cell bundles in the interstitium (D). **(E, F)** Patient III.4 (family 3), biopsy at 21-25 yrs. Advanced chronic renal injury (glomerulosclerosis, tubular atrophy, interstitial fibrosis) (E), interstitial smooth cell bundles (F). **(G, H – trichrome stain)** Patient III.1 (family 16), biopsy at 26-30 yrs. Tubular atrophy, moderate interstitial fibrosis. **(I, J – trichrome stain)** Patient III.4 (family 18), biopsy at 21-25 yrs. Glomerulosclerosis (I), tubular atrophy, severe interstitial fibrosis (J). Refer to Table 1 for further details.

Scale bars:

A, B – 100 µm

C, D, E, F, G, H, I, J – 200 µm

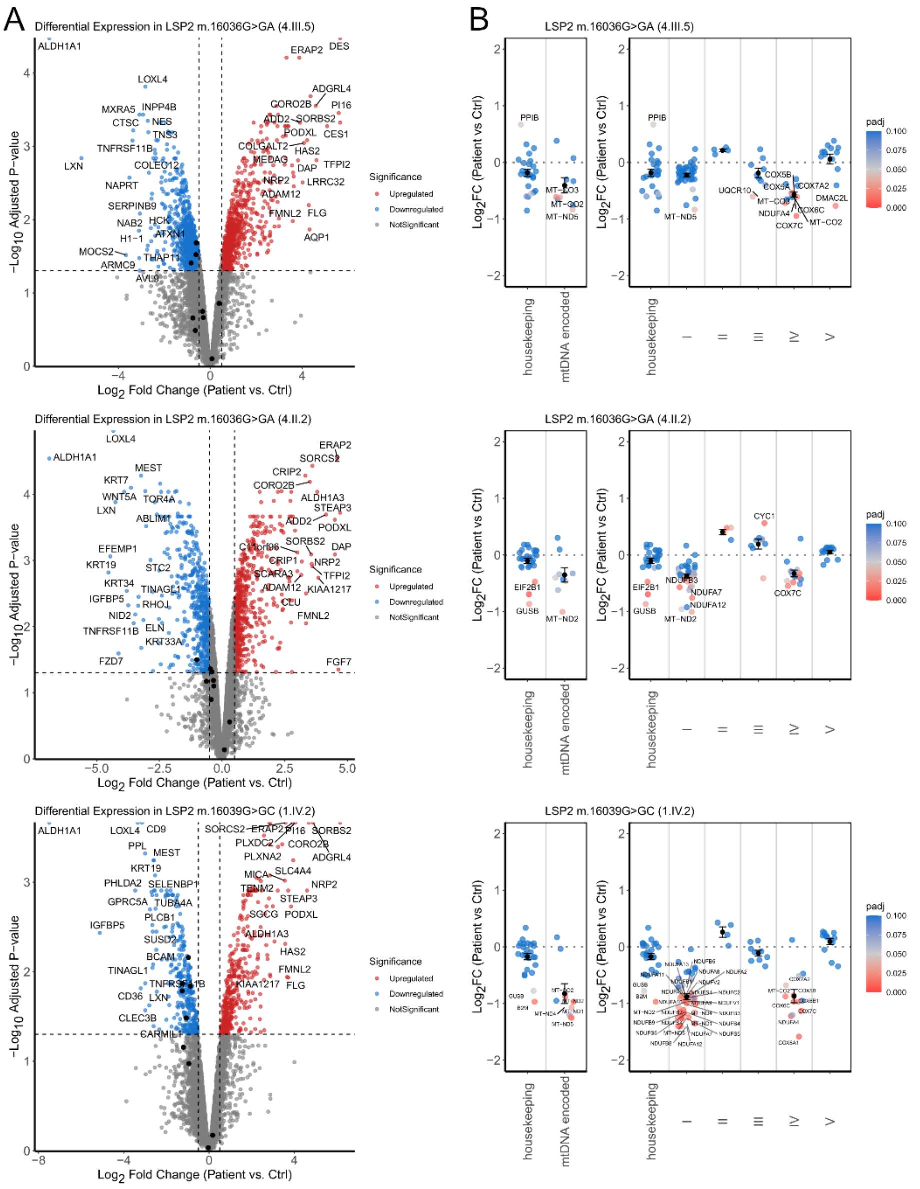


**Supplementary Figure S2: Non-targeted proteomic analysis of fibroblasts from patients with LSP2 (4.III.5, 4.II.2, 1.IV.2) variants. (A)** Volcano plots of all quantified proteins. Significantly upregulated proteins are shown in red, significantly downregulated proteins in blue, non-significant proteins in grey, mtDNA-encoded proteins are highlighted in black. **(B)** $Log_2$ fold change of mtDNA-encoded proteins and OXPHOS complex subunits. Black points indicate mean ± SEM.

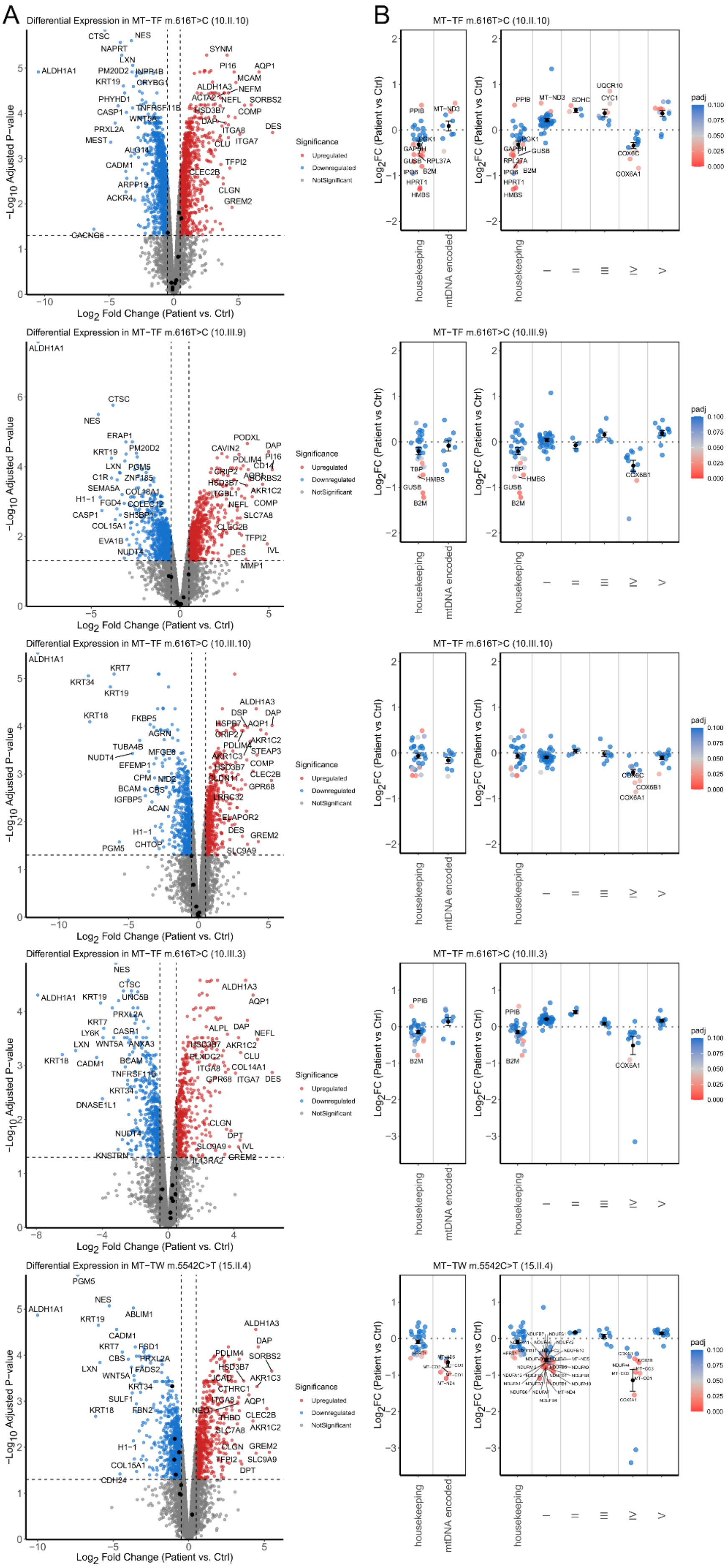
A
B
Differential Expression in MT−TF m.616T>C (10.II.10)
MT−TF m.616T>C (10.II.10)
Differential Expression in MT−TF m.616T>C (10.III.9)
MT−TF m.616T>C (10.III.9)
Differential Expression in MT−TF m.616T>C (10.III.10)
MT−TF m.616T>C (10.III.10)
Differential Expression in MT−TF m.616T>C (10.III.3)
MT−TF m.616T>C (10.III.3)
Differential Expression in MT−TW m.5542C>T (15.II.4)
MT−TW m.5542C>T (15.II.4)
−Log10 Adjusted P−value
Log2 Fold Change (Patient vs. Ctrl)
Log2FC (Patient vs Ctrl)
Significance
Upregulated
Downregulated
NotSignificant
padj
housekeeping
mtDNA encoded
I
II
III
IV
V

**Supplementary Figure S3: Non-targeted proteomic analysis of fibroblasts from patients with *MT-TF* (10.II.10, 10.III.9, 10.III.10, 10.III.3) and *MT-TW* (15.II.4) variants. (A)** Volcano plots of all quantified proteins. Significantly upregulated proteins are shown in red, significantly downregulated proteins in blue, non-significant proteins in grey, mtDNA-encoded proteins are highlighted in black. **(B)** $Log_2$ fold change of mtDNA-encoded proteins and OXPHOS complex subunits. Black points indicate mean ± SEM.

**Supplementary Table S1: Comparison of maternal vs. non-maternal clinical data from 224 adults identified by pedigrees.** Maternal are children of affected mothers, non-maternal are children of affected fathers.

| | Maternal | Non-Maternal | *P*-value (Z-Test for two proportions) |
|---|---|---|---|
| **Total Adults[1]** | **185** | **39** | |
| **Data available regarding development of ESRD (n)** | **185** | **39** | |
| ESRD = Yes | 45 (32%) | 0 | <0.00001 |
| ESRD = No | 140 (68%) | 39 (100%) | |
| Age of ESRD known | 36 | 0 | |
| Average Age ESRD | 47.6 ± 16.3 | -- | |
| **CKD history reported (n)[2]** | **119** | **17** | |
| CKD = Yes | 109 (92%) | 1 (6%) | < 0.00001 |
| CKD = No | 10 (8%) | 16 (94%) | |
| **Clinical data available (n)[2]** | **99** | **17** | |
| Clinically Affected = Yes | 89 (89%) | 1 (6%) | <0.00001 |
| Clinically Affected = No | 10 (11%) | 16 (94%) | |
| **Hyperuricemia/gout status available (n)** | **81** | **7** | |
| Hyperuricemia/Gout = Yes | 55 (63%) | 1 (17%)[3] | 0.0047 |
| Hyperuricemia and Gout = No | 26 (37%) | 6 (83%) | |
| Age of 1st gout attack known | 25 | 0 | |
| Average Age 1st Gout Attack | 34.9 ± 13.9 | -- | |
| **Hematuria status available (n)** | **41** | **5** | |
| Hematuria = Yes | 1 (2%) | 1 (20%) | 0.073 |
| Hematuria = No | 40 (98%) | 4 (80%) | |
| **Proteinuria status available (n)** | **36** | **4** | |
| Proteinuria = Yes | 5 (14%) | 0 | 0.426 |
| Proteinuria = No | 31 (86%) | 4 (100%) | |
| **Enuresis past age 4 status available (n)** | 45 | 6 | |
| Enuresis = Yes | 5 (11%) | 1 (9%) | 0.879 |
| Enuresis = No | 40 (89%) | 6 (91%) | |

[1]Data not including minors (Maternal n=7; Non-Maternal n=4)

[2]There is no statistical difference between the 109/119 CKD history reported by self or family member vs. 89/99 CKD clinically affected status based on clinical data (age, eGFR or age ESRD) (P=0.665499)

[3]Reported in medical record with hyperuricemia, however last serum uric acid level was 3.4 mg/dL.

**Supplementary Methods**

**Ethical approval**

This study was approved by the Institutional Review Boards of the First Faculty of Medicine, Charles University in Prague and the Wake Forest University Health Science Institutional Review Board. It adhered to the Declaration of Helsinki. Patients provided informed consent.

**Clinical evaluation and study population**

The Wake Forest-Charles University Rare Inherited Kidney Disease Registry is comprised of over 1400 families referred to A.J.B. by physicians and/or family members since 1996.[1, 2] A systematic review of all families with inherited tubulointerstitial kidney disease was undertaken to evaluate for mitochondrial inheritance.

**Genetic investigations**

Genomic DNA of all available individuals was extracted from whole blood, hair follicles, buccal swabs or patient-derived fibroblasts using standard procedures. Whole-exome sequencing was performed on individually barcoded DNA samples using the KAPA HyperExome v2 kit (Roche) and the KAPA HyperChoice MAX 0.5 Mb T1 human mitochondrial panel (96 reactions; Roche, cat. no. 1000005603). Whole-genome PCR-free sequencing libraries were prepared using the KAPA EvoPrep kit (Roche). DNA libraries were sequenced on the NovaSeq X Plus platform (Illumina) using 150-bp paired-end reads to achieve 30×, n×, and 5× coverage for whole-exome, whole-genome, and low-coverage whole-genome sequencing, respectively.

Raw sequencing reads were subjected to quality control and filtering using fastp v1.0. High-quality reads were subsequently aligned to the hg19 reference genome, and the revised Cambridge Reference Sequence (rCRS) for mitochondrial DNA, (NC_012920.1), using Novoalign v4.04.03. Sorting, duplicate removal, and read group assignment were performed using Picard Tools v2.23.5. Mitochondrial variant calling was performed with FreeBayes v1.3.10 using parameters optimized for mitochondrial DNA analysis (-F 0.04 -m 20 -q 15 -C 5), enabling detection of low-frequency variants while maintaining high-confidence variant calls. Identified variants were annotated using Mitomap, and mitochondrial haplotypes were assigned using HaploGrep 3 v3.22 based on PhyloTree build 17. Heteroplasmy levels were determined as the proportion of mitochondrial reads carrying the variant relative to the total number of reads, using either quantitative PCR or NGS data.

For pedigree construction and clinical description, the following terminology was used: genetically affected individuals were genetically tested and confirmed to carry the variant; obligate at-risk carriers were untested relatives presumed to carry the variant based on maternal inheritance; and clinically affected individuals had an estimated glomerular filtration rate (eGFR) more than 2 standard deviations below the age- and sex-adjusted means.[3] Individuals with unknown clinical status had no available serum creatinine measurements. P-values for maternal inheritance were calculated using a two proportion Z-test.

**Molecular and structural correlates of identified LSP2 and tRNA variants**

Variants in LSP2 were mapped to the rCRS and analyzed in the context of predicted binding sites of core mitochondrial DNA transcription machinery components, including mitochondrial transcription factor A (TFAM), mitochondrial transcription factor B2 (TFB2M), and mitochondrial RNA polymerase (POLRMT).[4] Conservation of LSP spacer sequence lengths[4] was assessed across humans and three primate species retrieved from GenBank using the ClustalW program.[5] Disease-associated variants in the mitochondrial tRNA genes (*MT-TF*, *MT-TW* and *MT-TL2*) were mapped onto their predicted secondary structures, which were retrieved from MitoVisualize.[6] The positions of post-transcriptional modifications of the corresponding tRNAs were obtained from Suzuki et. al.[7]

***In vitro* transcription of synthetic promoters**

Synthetic DNA templates containing either single (HSP or LSP2) or dual (HSP and LSP2) human mitochondrial promoters were cloned into the pEX-A128 backbone. All constructs were inserted between HindIII and BamHI restriction sites, with the mtDNA fragment positioned between these sites. Single-promoter constructs contained either the HSP region (nt 486–741) or the LSP2 region (nt 15956–16105). The dual-promoter construct was generated by fusing a synthetic LSP2 fragment (nt 16105–15956) with an HSP fragment corresponding to nt 486–649, preserving their native relative orientation, and cloned into the same pEX-A128 backbone.

Constructs corresponding to patient-derived sequence variants containing single-nucleotide insertions (T or G) were prepared similarly. Additional dual-promoter templates carrying −2, −1, wild-type, +1, +2, +3, +4, or +5 nt insertions in either the LSP2 or HSP promoter were generated to assess spacing effects between promoter elements and the TFAM-binding site.

Linear DNA templates for transcription assays were generated by PCR amplification from these plasmids using primer pairs annealing within the vector backbone and extending into the mtDNA insert to produce defined run-off transcripts. The HSP single-promoter template generated a run-off transcript of 199 nt, whereas the LSP2 single-promoter template generated a run-off transcript of 183 nt. For all dual-promoter templates, transcription from HSP and LSP2 generated run-off products of 182 nt and 234 nt, respectively.

PCR products were purified using standard silica column methods prior to use.

*In vitro* transcription assays were performed as described previously[8, 9] with minor modifications. Reactions were carried out in transcription buffer containing 25 mM Tris-HCl (pH 8.0), 10 mM $MgCl_2$, 1 mM DTT, 0.1 mg/mL BSA, and RNase inhibitor. Standard reactions (25 µL) contained 4 nM DNA template, 20 nM POLRMT, 40 nM TFB2M, 200 nM TFAM, 400 µM ATP, 150 µM GTP, 150 µM CTP, 10 µM UTP, and α-$^{32}$P-UTP, unless otherwise stated. Reactions were incubated at 32°C for 30 min.

Reactions were terminated by addition of stop buffer containing proteinase K, followed by incubation at 42°C for 30 min. RNA products were ethanol precipitated, resuspended in formamide loading buffer, and separated on denaturing urea-polyacrylamide gels. Gels were analyzed by phosphorimaging.

For dual-promoter templates, transcription from LSP2 was quantified and normalized to transcription from HSP within the same reaction. One-way ANOVA was performed using GraphPad Prism to compare the means of three groups, assuming a Gaussian distribution, followed by Tukey's multiple comparisons test for pairwise comparisons

**Patient-derived materials**

Skin fibroblasts were maintained in Dulbecco's modified Eagle's medium (DMEM; P04-04510, PAN Biotech) supplemented with 10% heat-inactivated fetal bovine serum (iFBS; SV30160.03, GE Healthcare HyClone) and 1× antibiotic/antimycotic solution (XC-A4110, Biosera) at 37 °C in a humidified atmosphere containing 5% $CO_2$. Cells were cultured to 80–90% confluence prior to experiments. Cells were harvested using 0.05% trypsin/0.02% EDTA for 5 min at 37 °C, washed, and centrifuged at 300 × g for 5 min at 24 °C. Unless otherwise stated, cell pellets were stored at −80 °C until analysis.

**Metabolic pulse–chase labeling of mtDNA-encoded proteins**

Proteins encoded by mtDNA were labeled using the $^{35}$S-Protein Labelling Mix (mixture of labelled methionine and cysteine; Revvity NEG072) essentially as previously described.[10] Briefly, cells were incubated for 15 min in methionine/cysteine-free DMEM (21013024, Gibco), followed by 15 min in the same medium containing either emetine (100 µg/mL) for pulse labeling or anisomycin (100 µg/mL) for chase labeling. $^{35}$S-Protein Labeling Mix (~100 µCi/mL) was then added, and cells were labeled for 1 or 3 h. Subsequently, 250 µM cold methionine and cysteine were added for 15 min, after which cells were washed with PBS containing 250 µM cold methionine/cysteine and then with PBS alone. Cells were either harvested immediately (pulse samples) or cultured for an additional 20 h in standard DMEM (392-0415, VWR) supplemented with 5% fetal bovine serum (chase samples). Harvesting was performed by trypsinization, and the cells were centrifuged at 600 x g and 4 °C for 5 minutes, with two washing steps in PBS. Samples were analyzed by SDS-PAGE directly after harvesting.

**SDS-PAGE and Western blot analysis**

Samples for SDS-PAGE were denatured for 20 min at 40 °C in a sample lysis buffer containing 50 mM Tris-HCl pH 7.0, 4% (w/v) SDS, 10% (v/v) glycerol, 0.02% (w/v) Coomassie Brilliant Blue R-250 (Serva Blue R, 35051) and 2% (v/v) 2-mercaptoethanol and separated on a 16% polyacrylamide gels (Hoefer SE600X) using the Tricine buffer system (Schagger 2006). Proteins were transferred onto polyvinylidene difluoride membranes (Immobilon FL, 0.45 µm; Merck) by semidry electroblotting (0.8 mA/cm$^2$, 1.5 h) using a Trans-Blot SD apparatus (Bio-Rad). Radioactive signals were detected using Storage Phosphor Screens BAS-IP MS 2025 (GE Healthcare) scanned on a Typhoon Imager (GE Healthcare). Following radioactive signal detection, the membranes were rehydrated in methanol, blocked for 1 h with 5% defatted milk in TBST0.1, and subjected to immunodetection using primary antibody against citrate synthase (ab129095, Abcam) and fluorescent secondary antibody (A10043, Thermo Fisher Scientific). Fluorescence signals were detected using the Odyssey CLx scanner (LI-COR Biosciences) and quantified with Image Lab software (Bio-Rad).

**Content of mitochondrial respiratory chain complexes**

For native electrophoresis, samples were prepared according to Fernandez-Vizarra.[11] Pellets containing $2x10^6$ cells were resuspended in 200 µL of PBS, mixed with 200 µL of digitonin solution (4 mg/mL) and incubated on ice for 10 min with occasional vortexing. After that, 1 mL of cold PBS+PIC (1:500, P8340, Merck) was added and samples were centrifuged at 10,000 x g for 5 min at 4 °C. Pellets were washed using 1 mL of cold PBS+PIC. The digitonized pellets were resuspended in 100 µL of Sample Buffer (1.5 M Aminocaproic Acid, 50 mM Bis-Tris, pH to 7.0) + PIC (1:500) + Benzonase nuclease (1:500, 70664-3, Merck). The membranes were solubilized by adding 10 µL of 10% n-dodecyl β-D-maltoside for 5 min on ice and centrifuged at 20,000 x g and 4 °C for 30 min. Supernatants were mixed with 10 µL of Blue-Native Sample Buffer (0.75 M Aminocaproic Acid, 50 mM Bis-Tris, 0.5 mM EDTA, 5% Serva Blue G, pH to 7.0) and stored -80 °C before the electrophoresis. Samples were analyzed by BNE using 4-13% polyacrylamide gradient mini gels (Mini-PROTEAN III, Bio-Rad) and the imidazole buffer system.[12] Proteins were transferred onto polyvinylidene difluoride membranes (Immobilon FL, 0.45 µm; Merck) by semidry electroblotting (0.8 mA/cm², 1.5 h) using a Trans-Blot SD apparatus (Bio-Rad). Membranes were blocked for 1 h with 5% defatted milk in TBST0.1 and subjected to immunodetection using primary antibodies against Complex I (NDUFA9, 20312-1-AP, Proteintech), Complex II (SDHA, 14865-1-AP, Proteintech), Complex III (CORE2, ab14745, Abcam), Complex IV (COX4, 11463-1-AP, Proteintech) and ATP synthase (F1-α, 66037-1-Ig, Proteintech), and fluorescent secondary antibodies (A10038 and A10043, Thermo Fisher Scientific). Fluorescence signals were detected using the Odyssey CLx scanner (LI-COR Biosciences) and quantified with Image Lab software (Bio-Rad).

**Mitochondrial respiration measurement**

The oxygen consumption rate of fibroblast cells was measured at 37 °C using the Oroboros Oxygraph-2k, following a substrate-uncoupler-inhibitor titration (SUIT) protocol,[13] modified for our specific conditions. Prior to each measurement, the oxygen sensors were calibrated in air-saturated medium corresponding to the specific conditions used in the subsequent experimental run. Protein concentration was measured, and 300-600 µg of cellular protein previously resuspended in PBS were placed into the Oxygraph-2k chamber with 2 mL of MiR05 medium. Cells were permeabilized with digitonin (0.05 mg per mg of protein). Substrates and inhibitors were used in the following order and concentrations: 10 mM pyruvate, 2 mM malate, 1 mM ADP, 10 mM glutamate, 10 mM succinate, 10 mM glycerol 3-phosphate (OXPHOS respiration), 500 nM oligomycin, 0,5 - 3 µM FCCP (ETC respiration), 1 µM rotenone, 10 mM malonate, 1 µM antimycin A, 2 mM ascorbate, 1 mM TMPD (COX respiration), and 0.5 mM KCN. Oxygen consumption was expressed in pmol oxygen.$s^{-1}$.mg $protein^{-1}$.

**Total coenzyme Q10 content**

Frozen pellets (fibroblasts) were homogenized according to Bartosova[14] and a cell homogenate was used for CoQ10 determination as described.[15] Protein concentration was analyzed by Lowry method.[16]

Citrate synthase (CS) activity was determined according to Srere.[17] Protein content and CS activity were used for the normalization of the Q10 value. Total CoQ10 content was analyzed by HPLC (HPLC 20 prominence system, Shimadzu, Japan) with UV detection according to Mosca.[18] All chemicals were purchased from Sigma-Aldrich (St. Louis, MO, USA).

**Label-free quantification mass spectrometry analysis**

Label-free quantification mass spectrometry analysis (LFQ-MS) of cell pellets was performed by the Proteomics Service Laboratory at the Institute of Physiology and the Institute of Molecular Genetics of the Czech Academy of Sciences following the SP4 no glass bead protocol.[19] Briefly, cellular pellets (100 µg of protein) were solubilized by 1% SDS in 100 mM triethylammonium bicarbonate (TEAB) buffer, reduced with 10 mM tris(2-carboxyethyl)phosphine (TCEP), and alkylated with 40 mM chloroacetamide (performed together at 95 °C for 10 min). Proteins were digested overnight at 37 °C with MS-grade trypsin at a 1:40 enzyme-to-protein ratio. The resulting peptides were desalted using C18 StageTips. Approximately 500 ng of peptide digest per sample was separated on a C18 column on nanoUHPLC Dionex Ultimate 3000 and analyzed in data-independent acquisition mode (DIA) on an Orbitrap Exploris 480 mass spectrometer equipped with a FAIMS unit. DIA MS Thermo raw files were processed in Spectronaut (v. 20.3, Biognosys) using directDIA mode and human proteome UP000005640_9606.fasta (UniProt release 2025_01) and default setting with Precursor and Protein Q-value and PEP cutoff set at 0.01. Protein group quantities (PG.Quantity, MS2 level) from Spectronaut's protein report were analyzed in Perseus software (version 2.1.5.0).[20]

**Supplementary References**


1. Bleyer AJ, Kidd K, Robins V, *et al.* Outcomes of patient self-referral for the diagnosis of several rare inherited kidney diseases. *Genet Med* 2020; **22:** 142–149.

2. Olinger E, Hofmann P, Kidd K, *et al.* Clinical and genetic spectra of autosomal dominant tubulointerstitial kidney disease due to mutations in UMOD and MUC1. *Kidney Int* 2020; **98:** 717–731.

3. Astley ME, Chesnaye NC, Hallan S, *et al.* Age- and sex-specific reference values of estimated glomerular filtration rate for European adults. *Kidney Int* 2025; **107:** 1076–1087.

4. Tan BG, Mutti CD, Shi Y, *et al.* The human mitochondrial genome contains a second light strand promoter. *Mol Cell* 2022; **82:** 3646–3660 e3649.

5. Thompson JD, Gibson TJ, Higgins DG. Multiple sequence alignment using ClustalW and ClustalX. *Curr Protoc Bioinformatics* 2002; **Chapter 2:** Unit 2 3.

6. Lake NJ, Zhou L, Xu J, *et al.* MitoVisualize: a resource for analysis of variants in human mitochondrial RNAs and DNA. *Bioinformatics* 2022; **38:** 2967–2969.

7. Suzuki T, Yashiro Y, Kikuchi I, *et al.* Complete chemical structures of human mitochondrial tRNAs. *Nat Commun* 2020; **11:** 4269.

8. Posse V, Hoberg E, Dierckx A, *et al.* The amino terminal extension of mammalian mitochondrial RNA polymerase ensures promoter specific transcription initiation. *Nucleic Acids Res* 2014; **42:** 3638–3647.

9. Posse V, Shahzad S, Falkenberg M, *et al.* TEFM is a potent stimulator of mitochondrial transcription elongation in vitro. *Nucleic Acids Res* 2015; **43:** 2615–2624.

10. Cunatova K, Reguera DP, Vrbacky M, *et al.* Loss of COX4I1 Leads to Combined Respiratory Chain Deficiency and Impaired Mitochondrial Protein Synthesis. *Cells* 2021; **10**.

11. Fernandez-Vizarra E, Zeviani M. Blue-Native Electrophoresis to Study the OXPHOS Complexes. *Methods Mol Biol* 2021; **2192:** 287–311.

12. Wittig I, Braun HP, Schagger H. Blue native PAGE. *Nat Protoc* 2006; **1:** 418–428.

13. Cunatova K, Vrbacky M, Knezu M, *et al.* The cytochrome c oxidase subunit COX6B1 is required for redox-sensitive early assembly and late stabilization of complex IV. *J Biol Chem* 2026; **302:** 111070.

14. Bartosova T, Klempir J, Hansikova H. Coenzyme Q10: A Biomarker in the Differential Diagnosis of Parkinsonian Syndromes. *Antioxidants (Basel)* 2023; **12**.

15. Krizova J, Hulkova M, Capek V, *et al.* Microarray and qPCR Analysis of Mitochondrial Metabolism Activation during Prenatal and Early Postnatal Development in Rats and Humans with Emphasis on CoQ(10) Biosynthesis. *Biology (Basel)* 2021; **10**.

16. Lowry OH, Rosebrough NJ, Farr AL, *et al.* Protein measurement with the Folin phenol reagent. *J Biol Chem* 1951; **193:** 265–275.

17. Srere PA. [1] Citrate synthase: [EC 4.1.3.7. Citrate oxaloacetate-lyase (CoA-acetylating)]. *Methods in Enzymology*, vol. 13. Academic Press, 1969, pp 3–11.

18. Mosca F, Fattorini D, Bompadre S, *et al.* Assay of coenzyme Q(10) in plasma by a single dilution step. *Anal Biochem* 2002; **305:** 49–54.

19. Alan L, Opletalova B, Hayat H, *et al.* Mitochondrial metabolism and hypoxic signaling in differentiated human cardiomyocyte AC16 cell line. *Am J Physiol Cell Physiol* 2025; **328:** C1571–C1585.

20. Tyanova S, Temu T, Sinitcyn P, *et al.* The Perseus computational platform for comprehensive analysis of (prote)omics data. *Nat Methods* 2016; **13:** 731–740.
